# Targeting Cortico-Striatal-Amygdalar Networks via Theta-Band Frontoparietal Synchronization in Opioid Use Disorder: A Randomized tACS-fMRI Trial

**DOI:** 10.64898/2026.02.10.26346048

**Authors:** Ghazaleh Soleimani, Rayus Kuplicki, Martin P Paulus, Hamed Ekhtiari

## Abstract

**Background:** Theta-band oscillation is integral to fronto-parietal connectivity in the executive control network and its top-down regulation on subcortical areas. External frontoparietal synchronization using theta-frequency transcranial alternating current (tACS) is a technology to potentially engage this network. In this pre-registered, triple-blind, sham-controlled trial (NCT03907644), we tested this intervention targeting the right frontoparietal network in people with opioid use disorder (OUD) to measure network engagement and behavioral outcomes.

**Method:** Sixty male participants with OUD were randomized to receive 20 minutes of active or sham 6 Hz tACS (HD electrodes over F4 and P4). Structural, resting-state, task-based fMRI drug cue reactivity, and repeated cue-induced craving assessments were collected immediately before and after stimulation. Pre-registered outcome measures were analyzed using time×group interaction models to examine (1) modulation of drug cue–related brain activity, (2) changes in craving, (3) alterations in functional connectivity, and (4) relationship between electric field, neural responses, and craving behavior.

**Results:** (1) A significant Time × Group interaction revealed decreased post-stimulation opioid cue–related activity in the active group relative to sham, involving key nodes in reward processing (ventral striatum, amygdala and ventral tegmental area) (FWE corrected α=0.05) (2) subjective craving did not differ significantly between groups (3) Group by time generalized psychophysiological interaction analyses showed increased right frontoparietal network engagement (β=2.63, p=0.0308) following stimulation, and increased top-down inhibitory regulation of frontoparietal network on right ventral striatum (β=1.99, p=0.037) and left medial amygdala (β=1.97, p=0.039) (4) Electric field strength in the right frontal/parietal node predicted frontoparietal network engagement in the active group (r=0.43, p=0.02).

**Conclusion:** Together, these findings demonstrate that theta-band frontoparietal tACS can modulate activity and task-dependent coupling within cortical–subcortical circuits in OUD, supporting network-targeted neuromodulation as a potential intervention for addiction.

**Significance Statement:** Addiction is linked to imbalances in cortico-subcortical brain circuits that control reward processing and craving. This study tested whether a non-invasive brain stimulation method— theta-band transcranial alternating current stimulation (tACS)—can rebalance these circuits in people with opioid use disorder. Using advanced brain imaging, we found that tACS strengthened communication within frontoparietal brain regions involved in self-control while reducing their connections with reward and emotion centers. These brain changes were linked to reduced craving responses to drug cues. Our results demonstrate that dual-site, network-targeted tACS modulates neural activity and task-dependent engagement of brain circuits during drug cue reactivity in addiction, supporting its potential as a novel therapeutic approach.

## Introduction

Opioid use disorder (OUD) is a chronic health condition affecting millions of individuals worldwide [1]. It is associated with increased vulnerability to trauma, legal complications, and premature mortality [2, 3]. Neuroimaging research, particularly functional MRI (fMRI) drug cue reactivity (FDCR), has substantially advanced our understanding of substance use disorders (SUDs) in general and OUD in particular [4, 5]. For example, in individuals with OUD, baseline FDCR in the nucleus accumbens, orbitofrontal cortex, and amygdala was significantly associated with addiction severity, and the relationship between nucleus accumbens FDCR and drug use severity was mediated by withdrawal symptoms [6].

Beyond regional effects, FDCR studies provide evidence for broader network-level disruptions [7]. Drug cue processing in individuals with addiction engages neural circuits that overlap with reward and emotion processing systems, including the striatum and amygdala [8]. There are also circuits involved in managing cravings and preventing relapse. Central to this process is top-down modulation by the executive control network, particularly frontoparietal regions such as the dorsolateral prefrontal cortex (DLPFC) and posterior parietal cortex (PPC), which exert regulatory control over subcortical structures, including the striatum and amygdala [9, 10].

Consistent with this network-level framework, FDCR paradigms provide a valuable basis for informing the development of interventions and evaluating treatment-related neural changes in SUDs [11, 12]. For example, interventions such as cognitive behavioral therapy [13], mindfulness-based approaches [14], and pharmacotherapies such as varenicline [15] have been shown to modulate cue-induced brain activation. Extending these insights, integrating treatment approaches that directly target addiction-related brain networks may further advance our understanding of the neural mechanisms underlying treatment response. In this context, the integration of noninvasive brain stimulation (NIBS) with FDCR paradigms offers an opportunity to modulate dysfunctional brain networks and assess neural responses to stimulation [16, 17].

In this regard, NIBS holds promise for targeting these networks to improve clinical outcomes in individuals with SUDs [16, 17]. Although there is compelling evidence supporting the use of transcranial magnetic stimulation (TMS), including FDA clearance of deep TMS for smoking cessation [18], electrical stimulation approaches still offer important advantages. Compared with TMS devices, transcranial electrical stimulation (tES) is relatively low-cost, portable, and scalable, showing promising effects in the field of addiction [19]. Furthermore, the recent FDA clearance of the first at-home tES device for depression further highlights the growing clinical feasibility of electrical stimulation approaches [20]. Among available tES techniques, alternating current stimulation (tACS) offers unique advantages for promoting network synchronization. Prior research suggests that effective communication between brain regions relies on the synchronized oscillatory activity of neurons [21, 22]. Although conventional electrical stimulation using large electrode pads induces relatively diffuse current flow compared with focal TMS, high-definition (HD) electrode configurations enable more focal modulation of underlying brain function using tES [23].

Dual-site HD-tACS has recently emerged as a novel form of NIBS, involving the placement of two sets of 4x1 HD electrodes over distinct cortical targets [24-30]. This approach has been utilized to influence endogenous neural oscillations between brain regions, thereby modulating cognitive functions. The effects of dual-site tACS on brain activity have been found to depend on the phase and frequency between stimulation sites. EEG recordings acquired during dual-site tACS on frontal and parietal nodes revealed that in-phase stimulation, where both sites are stimulated within the theta frequency band, enhanced synchronization between the targeted nodes and led to improvements in executive functions [24, 25]. Although neuroimaging studies have revealed disrupted frontoparietal synchrony and highlighted the role of its top-down regulation of the ventral striatum and amygdala in addiction [31, 32], and despite the potential of dual-site tACS to modulate this circuitry [33], no published studies have yet applied this approach to target frontoparietal synchronization in individuals with addiction.

To address this gap, we conducted the first pre-registered, randomized, triple-blind, sham-controlled clinical trial (NCT03907644) applying dual-site in-phase HD-tACS over the frontoparietal network in individuals with OUD. In this study, we investigated whether theta-band frontoparietal tACS can modulate neural activity and task-dependent engagement of brain circuits involved in cue reactivity and craving. Our pre-registered outcome measures included: (1) changes in subjective craving, (2) modulation of drug cue–related brain activity, (3) context-dependent changes in functional coupling within the frontoparietal network and between frontoparietal and subcortical regions, and (4) associations among electric field strength, neural responses, and craving. By using fMRI as an outcome measure, this study provides an opportunity to examine how an interventional manipulation of frontoparietal networks influences addiction-related brain function during cue reactivity. We hypothesized that theta-band frontoparietal tACS would modulate neural activity and task-dependent coupling within cortical–subcortical circuits engaged during cue exposure in addiction. This network engagement with a single-session intervention in an experimental trial can inform and optimize future larger-scale clinical trials with a higher number of sessions and longer follow-up.

## Method

### Participants

A total of 74 potential participants were screened for eligibility in this trial. Fourteen subjects withdrew or failed the screening process (see supplementary materials S1 for inclusion/exclusion criteria). The final randomization phase included sixty participants (all male, mean age ± SD=33.75 ± 7.04 years, ranging from 20 to 52) with opioid use disorder (OUD). All participants completed the imaging and stimulation sessions, contributing data for analysis. Recruitment took place from June 2019 to June 2023, during the participants’ early abstinence period at the 12& 12 residential drug addiction treatment center in Tulsa, Oklahoma. The trial protocol and primary and secondary outcomes were preregistered on ClinicalTrials.gov (Identifier: NCT03907644). Written informed consent was obtained from all participants before participation, and the study was approved by the Western IRB. This study was conducted in accordance with the Declaration of Helsinki, and all methods were carried out following relevant guidelines and regulations.

### Experiment Design

This randomized, triple-blind, sham-controlled trial included two parallel arms. Participants were randomly assigned to active or sham groups using computer-generated codes. To preserve blinding, group labels were coded as A and B. Data analysts conducted all analyses using these labels without knowledge of group identity. Unblinding occurred only after pre-registered data analysis for primary and secondary outcomes and the preparation of initial results. Exploratory analyses are performed after unblinding. MRI data, including structural MRI, resting-state fMRI, and fMRI drug cue reactivity tasks, were collected before and after each stimulation session. Behavioral data, including craving scores, were gathered at baseline, immediately before and after MRI scans, inside the scanner, and one day after the stimulation session. Adverse effects and blinding efficacy were assessed using structured questionnaires at two timepoints (t4 and t5; see Figure 1 and supplementary Tables S2–S3). Participants reported their guess of stimulation type, confidence (0–100), perceived craving reduction (Yes/No), and effectiveness (0–100). They also reported any side effects (e.g., headache, tingling), rated their severity, and judged their relation to tACS.

**Figure 1.**
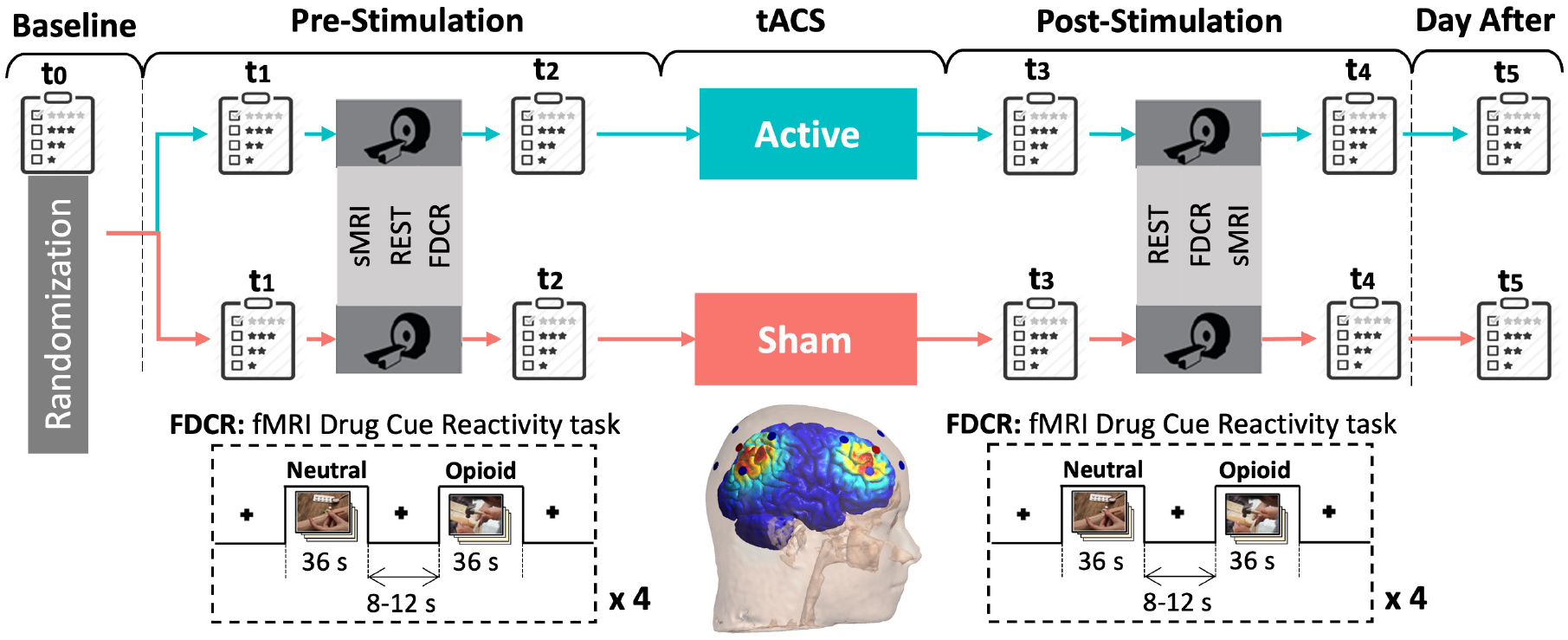
Experiment Design. In this randomized, triple-blind, sham-controlled clinical trial, 60 participants with opioid use disorders were randomly assigned to receive 20 minutes of active or sham 6 Hz tACS over the right frontoparietal network, using two sets of 4x1 HD electrodes placed over F4 and P4 locations in the EEG standard system. MRI data, including structural MRI (sMRI), resting-state fMRI (REST), and fMRI drug cue reactivity (FDCR) data using a standard block-designed task, were collected before and after stimulation. Self-report data, including craving visual analogue scale (VAS 0-100) scores, were collected at six different time points from t0 (baseline) to t5 (day after stimulation).

### Outcomes

The **primary outcome** focuses on the change in blood oxygen level–dependent (BOLD) signal during a drug cue exposure fMRI task, specifically examining differences in activation across regions of interest, including the prefrontal cortex, insula, striatum, and extended amygdala, comparing drug-related versus neutral stimuli. The **secondary outcomes** are assessed changes in self-reported drug craving, measured via a visual analog scale (0–100), and alterations in functional connectivity between subcortical regions and cortical areas such as the prefrontal cortex and insula. **Exploratory outcomes** investigate additional measures that examine the interplay between neural activity and connectivity, subjective craving, and electric field.

### Data Collection

#### Intervention

Two sets of 4×1 HD electrodes targeted the right dorsolateral prefrontal cortex (centered at F4) and right inferior parietal cortex (centered at P4). Active stimulation involved 20 minutes of in-phase 6 Hz frontoparietal tACS at 2 mA peak-to-peak over F4 and P4, with 0.5 mA return currents (180° out of phase) at surrounding electrodes. The right frontoparietal network was selected due to its dominant contribution among individuals with SUD, and the right FPS has been reported to have more pronounced effects on executive functions. In the sham condition, the device applied brief stimulation at the beginning and end (30-second ramp-up and ramp-down) to mimic the tingling sensation of real stimulation (Figure 2 and supplementary materials S3 for more details on stimulation parameters and randomization).

**Figure 2.**
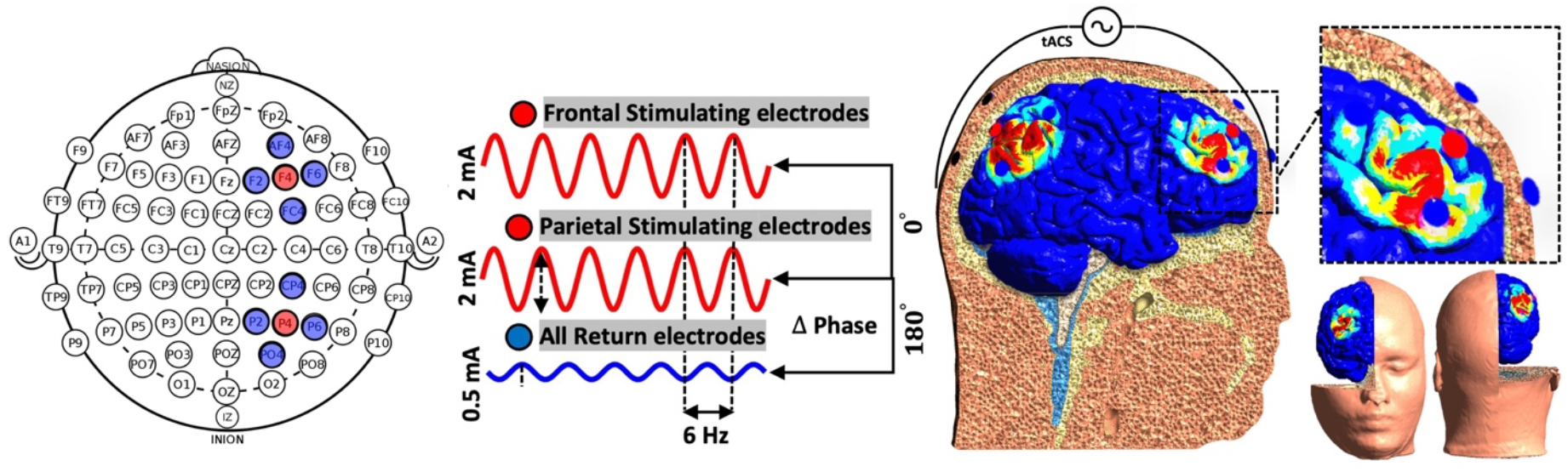
Schematic of 6 Hz HD-tACS Montage and Modeled Electric Field Distribution. **Left:** EEG topographic map illustrating electrode placement with frontal (F4) and parietal (P4) stimulating electrodes (red), and surrounding return electrodes (blue). **Center:** The stimulation waveform design consists of in-phase sinusoidal currents at 6 Hz with 2 mA peak-to-peak amplitude applied to both frontal and parietal sites, and 0.5 mA distributed to the return electrodes. **Right:** Finite element model of electric field distribution for one sample subject.

### MRI Data

Resting-state fMRI was acquired in 8-minute runs immediately before and after stimulation, followed by a 6.5-minute block-design drug cue reactivity task. During rest, participants fixated on a cross and cleared their minds. The task included alternating blocks of opioid-related and neutral images (6 images per block, 5 seconds each, with 0.2-second inter-stimulus intervals; as validated here [34]), interleaved with 8–12 second fixation intervals. Craving ratings were collected after each block (see Supplementary Materials S4–S5 for MRI parameters and task details).

### Electric-Field Modeling

To quantify individual variability in stimulation dose, we performed subject-specific electric field simulations using computational head models. These analyses estimated the spatial distribution and magnitude of the induced electric field in each participant and assessed whether individual differences in electric field strength were associated with stimulation-related neural and behavioral changes. For each participant, the 99th percentile of the electric field within the frontal and parietal cortices was identified and transformed into MNI space. Spherical ROIs (10 mm radius) were then centered on these coordinates to standardize ROI definition while preserving individual variability in targeting condition (see Supplementary Sections S6.2 and S6.3 for details). Average electric-field strength extracted from these individualized ROIs was entered into correlation and regression models together with stimulation condition (active vs. sham) to test whether electric field–behavior and electric field–neural relationships differed by stimulation.

### Statistical Analysis

#### Sample size

The preprocessed sample size of 30 participants per arm (see Table 1 for demographic data in baseline assessments) provided 80% power to detect an effect size (Cohen’s d) of 0.74 for changes in drug-cue reactivity between the arms at a two-sided 0.05 significance level in a two-sample t-test.

**Table 1.**
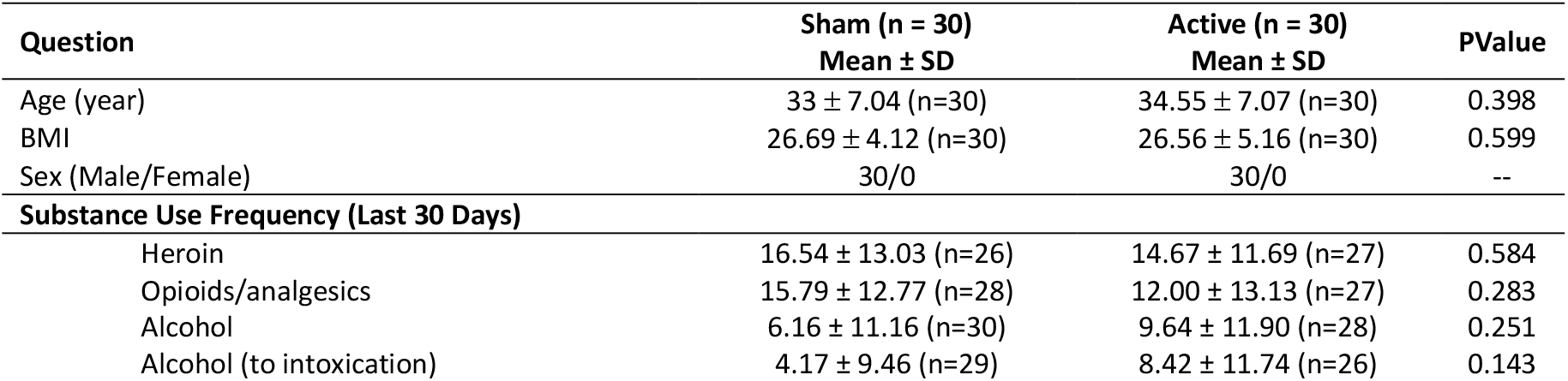
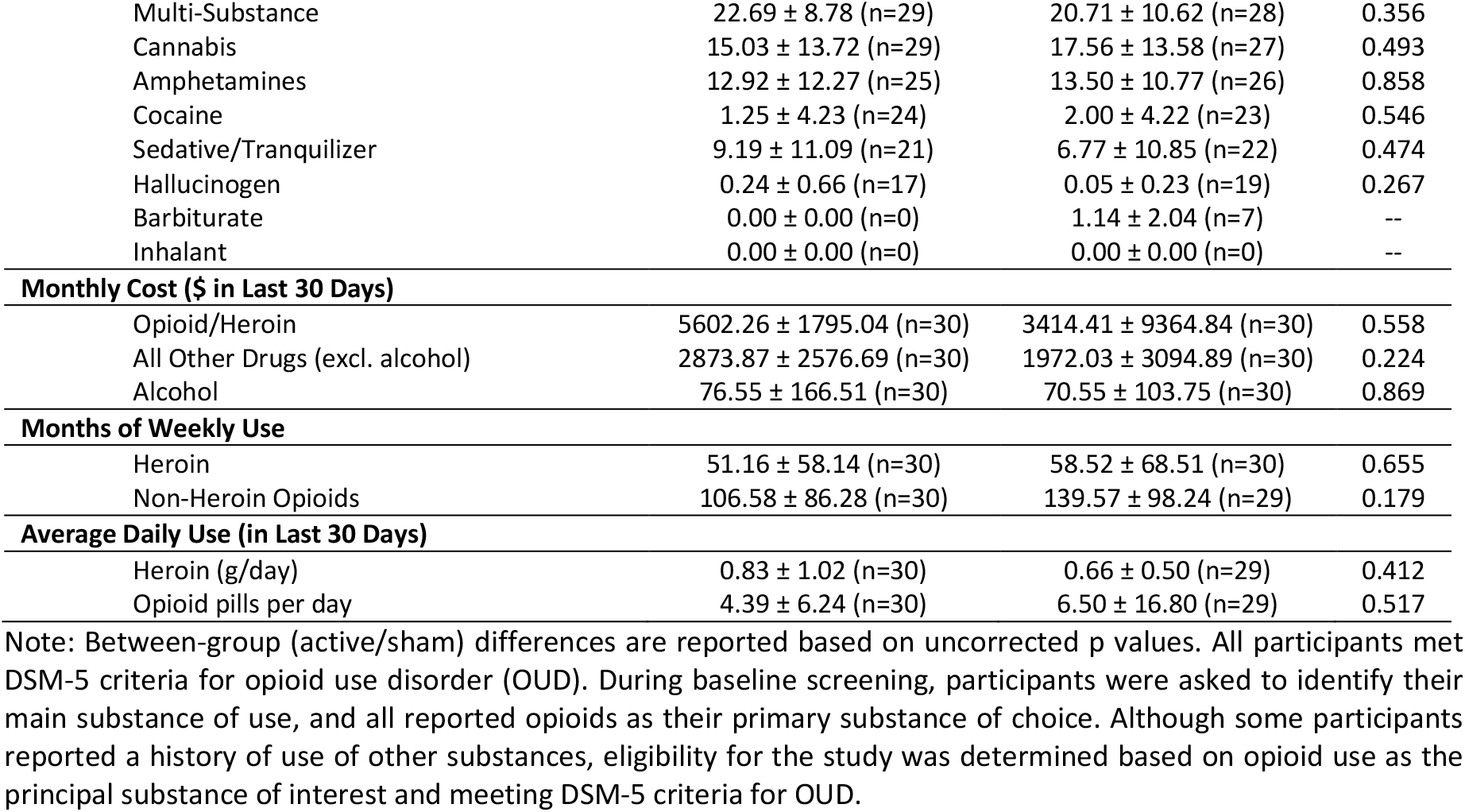
Demographic Data.

In the following sections, model assumptions, including normality and homogeneity of variance, were assessed using residual diagnostics and Levene’s tests where applicable.

#### Primary hypothesis

We hypothesized that active frontoparietal synchronization would reduce cue reactivity in striatal and amygdala subregions compared to sham. Preprocessing details are provided in Supplementary S6. First-level block design analyses (opioid > neutral) were conducted using GLMs. Brainnetome atlas parcellation was applied, and beta values were extracted per subregion to visualize whole-brain activity over time. For voxelwise group-level analyses, AFNI’s 3dLME was used to fit linear mixed-effects (LME) models with Group (active vs. sham), Time (pre vs. post), and their interaction as fixed effects, and Subject as a random effect (-ranEff ‘∼1|Subj’). Statistical maps were thresholded at voxelwise p<0.005, and a cluster-extent threshold was determined using AFNI’s 3dClustSim (combined with 3dLocalACF for non-stationary cluster-level correction, 10,000 iterations, study-specific spatial autocorrelation function parameters) to achieve a family-wise error (FWE) corrected α=0.05. This procedure corresponded to clusters ≥40 contiguous voxels.

#### Secondary hypothesis (behavioral)

To check the first secondary hypothesis regarding the efficacy of frontoparietal stimulation on drug-related behaviors, craving scores, control of craving, and desire for drug questionnaires (DDQ) [35] were assessed at various time points before and after stimulation. Craving scores were also rated inside the scanner at the end of each block (more details in supplementary materials sections S2 and S5). Mean ± SD values were calculated for each group and time point. Group differences over time were evaluated using a mixed repeated-measure ANOVA with Group (active vs. sham) as a between-subjects factor and Time as a within-subjects factor, including the Group × Time interaction. When appropriate, post-hoc pairwise comparisons were performed (within- and/or between-group contrasts), and p-values were adjusted using FDR correction.

#### Secondary hypothesis (neural)

To examine the other secondary hypothesis, we tested whether frontoparietal stimulation increased interactions between frontal-parietal nodes and frontoparietal–subcortical connections. ROI-to-ROI resting-state and task-dependent (generalized psychophysiological interaction (gPPI)) connectivity were performed using the CONN toolbox. Resting-state connectivity was based on correlation analysis. The task conditions were also modeled using boxcar functions of the cue reactivity task convolved with a canonical hemodynamic response function (HRF). The design matrix for gPPI included the seed region’s time course (physiological term), task time course (psychological term), and the interaction between the task and BOLD signal in the seed region (psychophysiological (PPI) term). Covariates such as the BOLD signal of white matter and CSF, along with motion parameters, were used to remove unwanted motion and physiologic artifact effects for both resting-state and task-based connectivity. For the second-level analysis, the interaction of time (pre vs post) by group (active vs. sham) was calculated. For the ROI-to-ROI analysis, results were reported when FDR corrected P<0.05. Seed definition approaches, including frontoparietal seeds and subcortical regions like striatum and amygdala, can be found in supplementary materials S6. In sum, individual peak electric fields in the frontal and parietal cortices (underneath the stimulation sites and their contralateral counterparts) were mapped to MNI space, and 10 mm spheres were created at these locations (supplementary materials S6.3). The Brainnetome and Oxford-Harvard atlases were used to define the amygdala and striatal subregions, respectively. These regions served as the main nodes for cortico-subcortical connectivity analyses. Bar plots showing mean ± SD connectivity by group changes (e.g., right DLPFC–parietal) were added (Figure 4) to make the effects more interpretable. Electric field strength, which was significantly correlated with frontoparietal PPI changes in the active group, was included as a covariate in regression models examining PPI–craving associations to disentangle the effects of stimulation from connectivity.

#### Exploratory hypotheses

In addition, we pre-registered an exploratory hypothesis testing whether individual differences in tACS-induced electric field strength were associated with changes in functional activity, frontoparietal connectivity, and craving. To evaluate this hypothesis, we conducted correlation and regression analyses examining the relationships between electric field strength, neural activity/connectivity changes, and craving scores in both the active and sham stimulation groups.

## Results

Demographic and substance use characteristics for the active and sham groups are presented in Table 1 (and supplementary materials section S7). Blinding was generally effective, with most participants guessing they had received real stimulation (Sham/Active: 24/25 post-fMRI [t4], 27/23 the day after [t5]). Only one participant in the sham group reported a severe adverse effect (trouble concentrating) immediately after stimulation, and one participant in the active group reported severe skin redness the following day. No other severe effects were reported (see supplementary section S8 for further results on blinding and adverse effects).

Homogeneity of variance was assessed using Levene’s tests and residual diagnostics, and no significant between-group differences in variance were detected for the analyzed outcomes.

### Primary outcome

Whole-brain LME analysis of functional activity data for the opioid vs. neutral contrast revealed the largest significant cluster with a time x group interaction over the bilateral striatum, amygdala, ventral tegmental area (VTA), and posterior cingulate cortex (PCC), consisting of 864 voxels with peak activation at [19 -35 7] in MNI space. This cluster with main nodes in the reward network and PCC in the default mode network exhibited a significant decrease in functional activity in response to opioid vs. neutral cues after stimulation compared to before stimulation in the active group, relative to the sham group (Figure 3).

**Figure 3.**
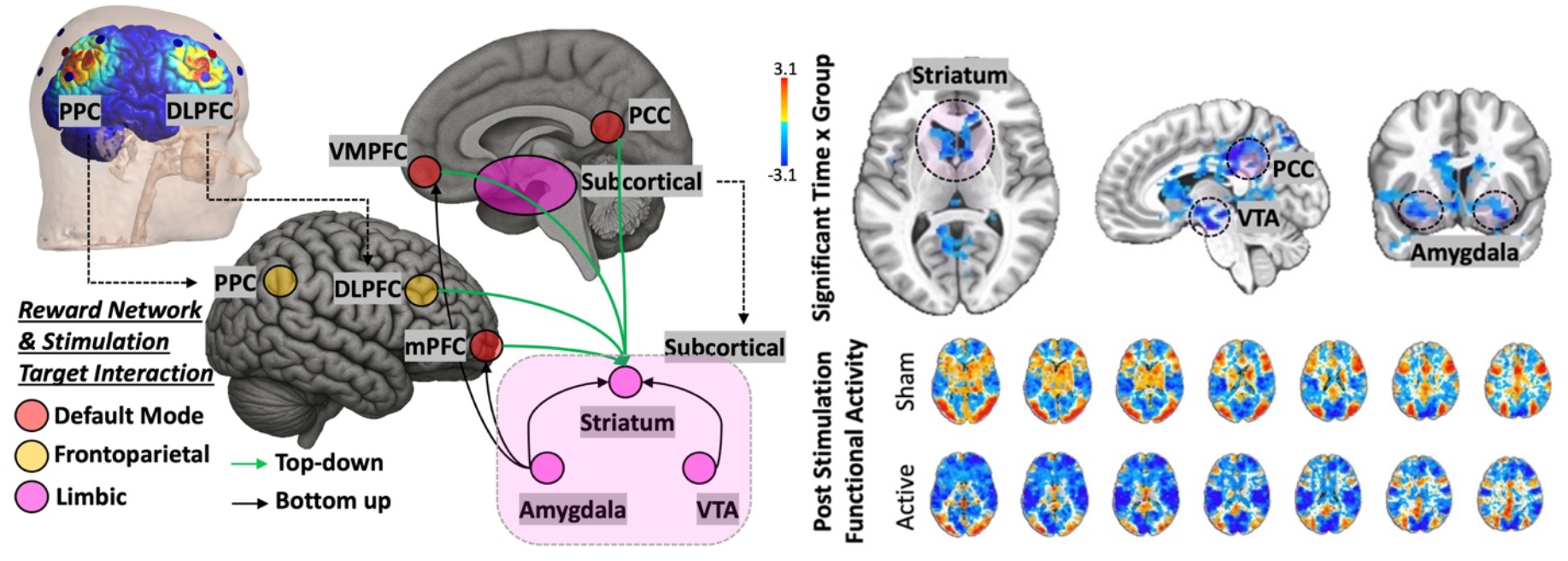
Whole Brain Functional Activity Analysis Results. The left panel shows the hypothesis behind cortico-subcortical interaction through the reward system and large-scale brain networks. In the right panel, the upper panel displays the FEW-corrected Z map obtained from LME analysis. The lower part shows the uncorrected post-stimulation map for active and sham groups. <u>Abbreviations:</u> DLPC: dorsolateral prefrontal cortex, PPC: posterior parietal cortex, mPFC: medial prefrontal cortex, VMPFC: ventromedial prefrontal cortex, ACC: anterior cingulate cortex, PCC: posterior cingulate cortex, VTA: ventral tegmental area.

### Secondary outcome (behavioral)

The ANOVA results revealed significant effects of time on single-item craving VAS (0-100) scores (F(5, 344) = 14.21, p < 0.0001), indicating fluctuations over different assessment time points. However, the effect of group (active vs. sham) on craving VAS was not significant (F(1, 344)=0.056, p=0.8133), nor was there a significant interaction between time and group (F(5, 344)=0.386, p=0.8581). Similar results were obtained for the time x group interaction for DDQ total score. However, post hoc analysis showed a trend toward a significantly lower DDQ desire for drugs sub score in the active group (1.68 ± 0.82) compared to the sham group (1.34 ± 0.5) (p=0.0613), day after and also immediately after post fMRI (sham: 1.92 ± 1.03, active: 1.49 ± 0.81, p=0.071). (See supplementary materials S9)

### Secondary outcome (neural)

To evaluate the effects of frontoparietal stimulation on functional connectivity during drug cue exposure, ROI-to-ROI generalized PPI analyses were conducted, using the right frontal (F4) or right parietal (P4) region, corresponding to the primary stimulation targets, as the seed. Homologous left frontoparietal cortical regions, along with striatal and amygdalar subregions, were included as target nodes to assess both ipsilateral and contralateral network-level effects of stimulation. Functional connectivity effects were evaluated using time (post vs. pre) × group (active vs. sham) interactions, which capture relative stimulation-induced changes over time between the two groups. An interaction was considered an increase in PPI connectivity when group-level changes in PPI beta values (post − pre) were greater in the active group compared with the sham group (shown in red in Figure 4). In contrast, an interaction was considered a **decrease in PPI connectivity** when group-level changes in PPI beta values (post − pre) were lower in the active group relative to sham (shown in blue in Figure 4). This definition and color-coding scheme follow standard conventions used in the CONN toolbox.

For ROI-to-ROI analyses, (a) the right frontal or (b) the right parietal node was used as the seed. For each seed, three sets of target regions were examined: (1) other frontoparietal nodes as targets, (2) striatal subregions added to the frontoparietal targets, and (3) amygdala subregions added to the frontoparietal targets. Significant effects in all three sets were observed only when the right parietal node was used as the seed, as detailed below.

**Figure 4.**
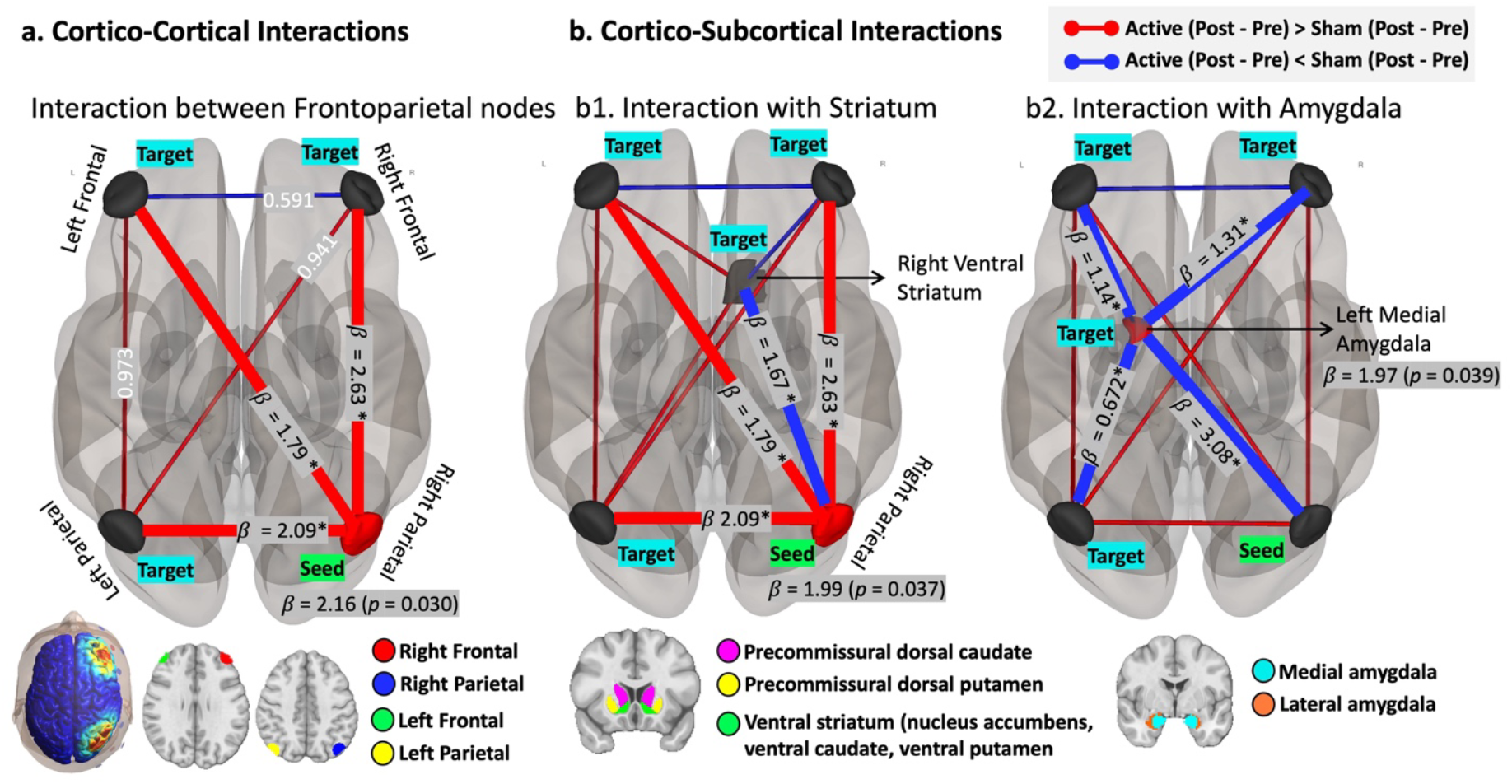
Time × group interaction effects on cue-related PPI connectivity. Task-dependent functional connectivity was assessed using generalized psychophysiological interaction (gPPI) analyses for the cue > neutral contrast, examining time × group (post vs. pre × active vs. sham) interactions. **<u>Red lines</u>** indicate greater post–pre connectivity changes in the active group relative to sham, whereas **<u>blue lines</u>** indicate lower post–pre connectivity changes in the active group relative to sham. **(a) Cortico-cortical interactions**. Using the right parietal stimulation site as the seed, and the right frontal, left frontal, and left parietal cortices as targets. The right parietal seed showed significantly higher post–pre connectivity with all other frontoparietal nodes in the active group relative to sham. **(b) Cortico-subcortical interactions. (b1) Striatal connectivit**y. Striatal subregions, including the left and right dorsal caudate, dorsal putamen, and ventral striatum, were added to the frontoparietal nodes shown in (a). The right parietal cortex was used as the seed, with the remaining frontoparietal regions and striatal subregions treated as targets. The right parietal seed showed significantly lower post–pre connectivity with the right ventral striatum in the active group compared with the sham group. **(b2) Amygdala connectivity**. Amygdala subregions, including the left and right medial and lateral amygdala, were added to the same frontoparietal seeds shown in (a). The right parietal cortex was used as the seed, with the remaining frontoparietal regions and amygdala subregions treated as targets. The left medial amygdala showed significantly lower post–pre connectivity with all four frontoparietal nodes in the active group relative to sham. Seed regions are highlighted in green and target regions in cyan. Line thickness reflects the magnitude of the interaction effect. Statistical values (β coefficients and FDR-corrected p-values) are shown for significant connections. The transparent head model illustrates the distribution of the tACS-induced electric field.

1. **Cortico-cortical interactions**. Using the right parietal cortex as the seed, significant time × group interaction effects were observed for connectivity with the right frontal, left frontal, and left parietal cortices (β = 2.16, p = 0.0308, FDR-corrected). These interactions reflected greater increases in gPPI values from pre-to post-assessment in the active stimulation group relative to the sham group for connections with the right frontal (β = 2.63), left frontal (β = 1.79), and left parietal (β = 2.09) regions.
2. **Cortico-striatal interactions**. When striatal subregions were added to the frontoparietal targets, a similar interaction cluster emerged in the cortico-striatal analysis (β = 1.99, p = 0.037, FDR-corrected). In this case, the time × group interaction reflected a greater reduction in gPPI values from pre-to post-assessment in the active group compared with the sham group for connectivity between the right parietal cortex and the right ventral striatum (β = 1.67).
3. **Cortico-amygdala interactions**. When amygdala subregions were included as targets, a significant interaction was observed between the right parietal cortex and the left medial amygdala (β = 1.97, p = 0.039, FDR-corrected), indicating a greater pre-to post-assessment reduction in task-dependent coupling in the active group relative to the sham group. Exploratory interactions involving the insula are presented in Supplementary Section S9, and between-subject variability in post–pre connectivity values across frontoparietal nodes is shown in Supplementary Section S12 to further characterize the distribution and direction of the observed effects.

Exploratory interactions involving the insula are presented in Supplementary Section S9, and between-subject variability in post–pre connectivity values across frontoparietal nodes is shown in Supplementary Section S12.

For resting-state data, we observed increased functional connectivity at the stimulation sites (right frontal and parietal nodes), along with decreased connectivity between both left and right frontoparietal nodes and the left medial amygdala. However, these effects did not reach statistical significance due to limited power. Additionally, no significant connectivity changes were observed in striatal subregions (supplementary materials S10).

### Combined Brain-Behavior-Stimulation analyses

To explore the relationship between stimulation-induced electric fields, brain connectivity, and behavioral outcomes, we fit a linear regression model with cue-induced craving change (VAS: t4– t3) as the outcome and right frontoparietal PPI connectivity change (post–pre), stimulation group (active vs. sham), and their interaction as predictors. The group × connectivity interaction was significant (p=0.04), indicating that the relationship between frontoparietal connectivity and craving reduction differed by stimulation condition. Follow-up correlations revealed a significant negative association in the active group (r=–0.37, p=0.05), suggesting that greater increases in connectivity were associated with larger decreases in craving. No such association was found in the sham group (r=0.08, p=0.69), supporting the specificity of this brain–behavioral relationship to active stimulation (Figure 5). (See Supplementary Section S16 for further exploratory results).

**Figure 5.**
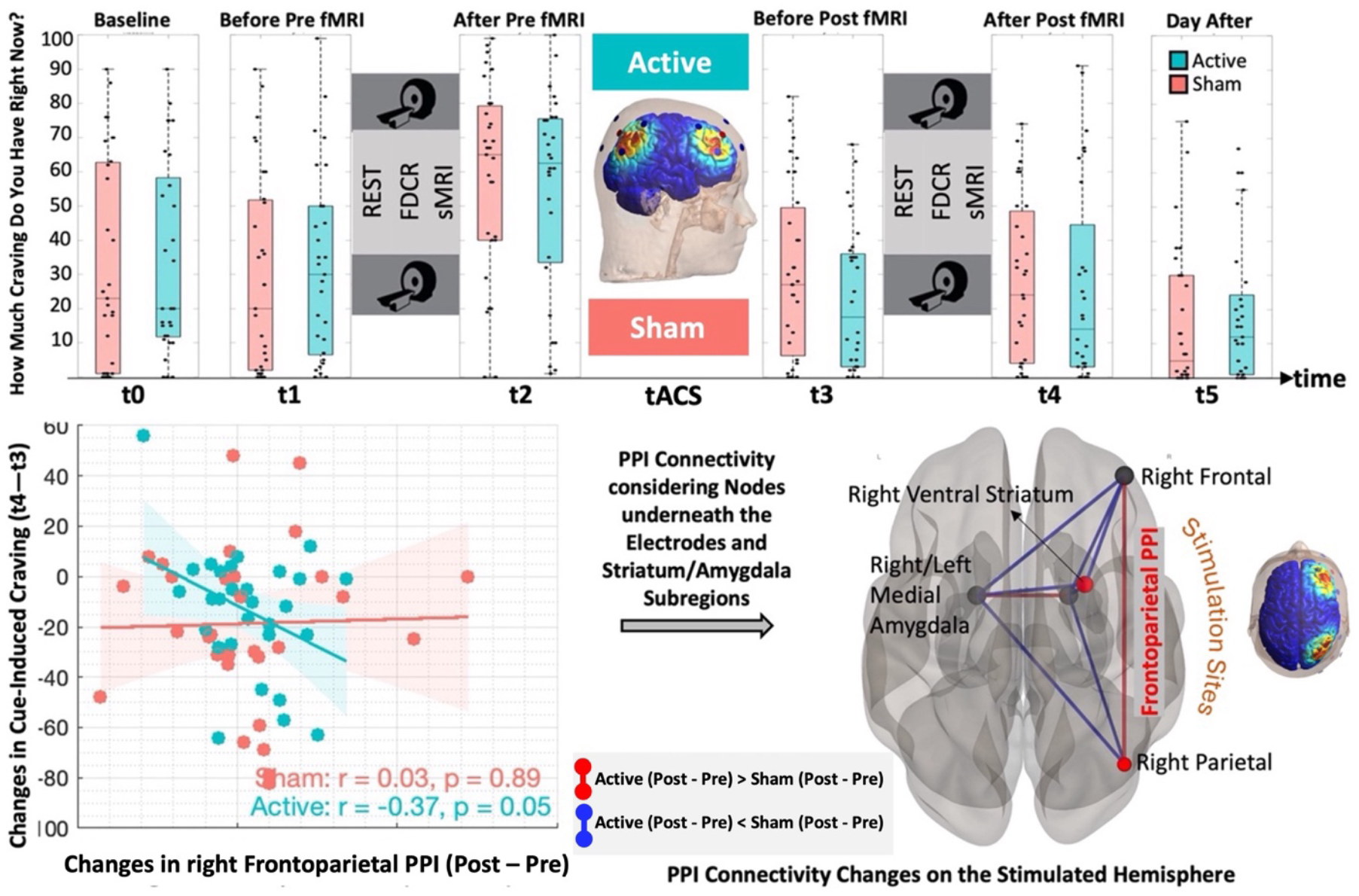
Effects of right frontoparietal tACS stimulation on craving and right frontoparietal connectivity. **Top:** Craving ratings across six time points—Baseline (t0), Before (t1)/After (t2) Pre-Stimulation-fMRI, Before (t3)/After (t4) Post-Stimulation-fMRI, and Day After (t5)—for Active and Sham conditions. Time points (t0 to t5) are defined based on the experimental design shown in Figure 1. **Bottom left:** Negative correlation between frontoparietal PPI changes and post-stimulation cue-induced craving changes (t4 – t3) in the Active condition (r = –0.37, p = 0.05), with no significant correlation in the Sham condition. Each dot represents one participant. **Bottom right:** Brain schematic highlighting significant frontoparietal PPI changes between the right frontal and right parietal regions, as well as limbic regions including the medial amygdala and ventral striatum. **<u>Red lines</u>** indicate greater post–pre connectivity changes in the active group relative to sham, whereas **<u>blue lines</u>** indicate lower post–pre connectivity changes in the active group relative to sham.

As illustrated in Figure 6, individualized electric field modeling revealed consistent targeting across participants. The group-averaged EF distribution map (left panel) showed focal peaks over both frontal and parietal regions, indicating reliable stimulation across the frontoparietal network. The left panel depicts the spatial distribution of each subject’s peak EF coordinate in the right frontal cortex, highlighting variability in peak field localization. To standardize region-of-interest (ROI) selection, 10 mm spheres were created around each individual’s peak coordinate (red). We fit a linear regression model with right frontoparietal PPI connectivity change (post–pre) as the outcome and stimulation group (active vs. sham), EF strength, and their interaction as predictors. The group × EF interaction was significant (p=0.03), indicating that the relationship between EF strength and PPI change differed by stimulation condition. Follow-up correlations revealed a significant positive association in the active group (r=0.43, p=0.02), but not in the sham group (r=0.08, p=0.69), consistent with a dose-dependent effect specific to active tACS.

**Figure 6.**
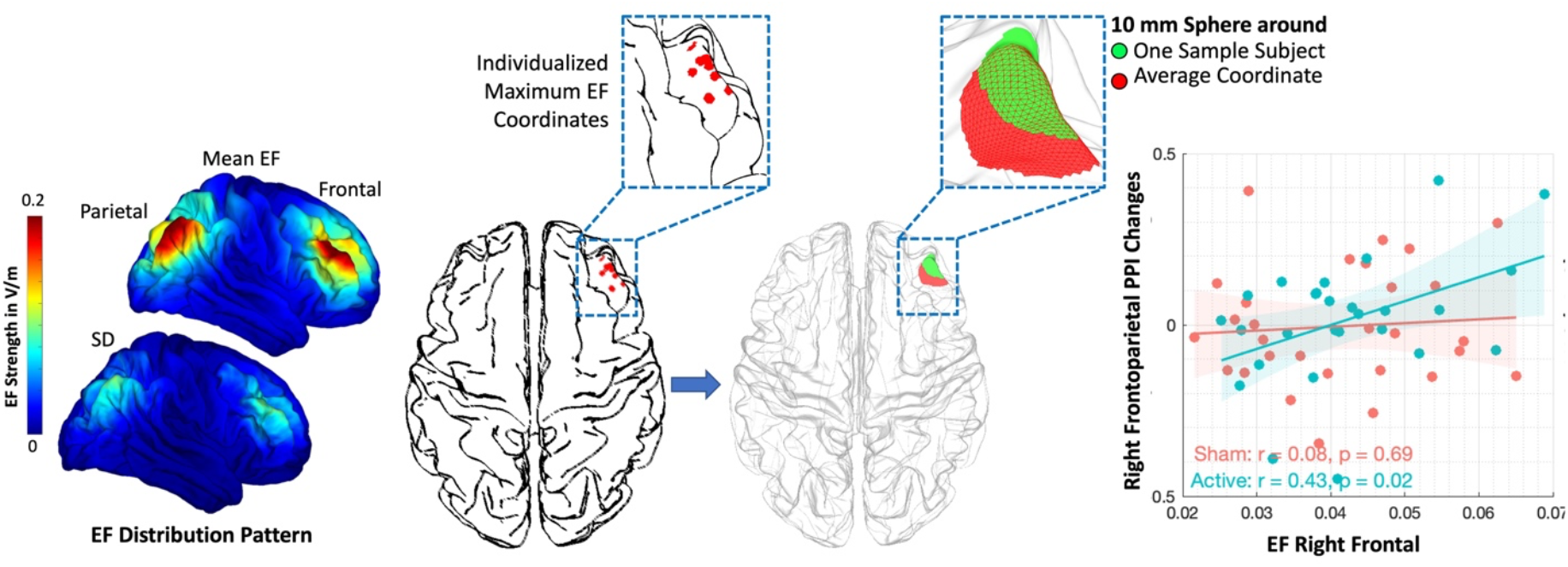
Relationship Between Electric Field (EF) Strength, Frontoparietal Connectivity, and Stimulation Targeting. The left panel illustrates individualized peak EF coordinates in the right frontal cortex. The second panel shows 10 mm spherical ROIs centered on one sample subject’s peak EF (red) and the group-level averaged coordinate (green). **Scatter Plot:** Correlation between EF strength in the right frontal cortex and frontoparietal PPI changes, shown separately for the Active (r = 0.43, p = 0.02) and Sham (r = 0.08, p = 0.69) conditions. **Colored Brain:** Group-level mean EF map and corresponding standard deviation (SD) map, highlighting the spatial consistency of stimulation across participants in both right frontal and parietal regions.

## Discussion

In this pre-registered HD-tACS-fMRI study, we investigated whether enhancing frontoparietal synchronization could modulate cortico-subcortical interactions in individuals with opioid use disorder (OUD). The study yielded four key findings: (1) <u>Primary outcome:</u> Linear mixed-effects models revealed a significant time x group interaction with a reduction in activations related to drug cue reactivity in brain areas related to reward and emotion processing, such as striatum, amygdala, VTA, and posterior cingulate cortex (PCC) in the active group compared to the sham group. (2) <u>Secondary outcome (behavioral):</u> Although cue-induced craving decreased in both active and sham groups following stimulation, no significant time × group interaction was observed for subjective craving using single item VAS (1-00). (3) <u>Secondary outcome (neural):</u> Task-based functional connectivity analyses showed significant time × group interactions, including increased frontoparietal PPI connectivity in both hemispheres and decreased PPI connectivity between the parietal cortex and ventral striatum, as well as between the frontoparietal network and medial amygdala, in the active versus sham group post-stimulation. (4) <u>Exploratory outcome:</u> Finally, increased frontoparietal connectivity during drug cue exposure was negatively correlated with cue-induced craving and positively correlated with electric field strength in the right frontal stimulation site.

Our results showed that craving increased significantly after the first cue exposure during pre-stimulation fMRI drug cue reactivity (FDCR) (t2 vs. t1), regardless of group. Following stimulation, both active and sham groups showed reduced craving (t3 vs. t2), but no further increase was observed after the second cue exposure (post-stimulation FDCR) (t4 vs. t3). Self-reported craving also declined from baseline to the day after stimulation in both groups, with no significant differences between them. Similarly, the desire for drugs assessed by the DDQ showed no significant group × time interaction. However, post hoc comparisons indicated a trend toward lower DDQ scores in the active group compared to the sham group, both immediately after stimulation (p=0.071) and on the following day (p=0.0613). Overall, while the combined effects of cue exposure and tACS reduced craving, no significant group-specific effects or time × group interactions were detected. Several single-session tES studies have similarly found no significant differences in self-reported drug craving between active and sham groups (e.g., Lupi et al., 2017, systematic review [36]). Although no significant group differences in subjective craving were observed, significant neural effects suggest that a greater number of stimulation sessions may be needed to produce greater behavioral changes, such as reduced craving, particularly in the active group.

Our primary outcome analysis revealed a significant time × group interaction in fMRI cue reactivity, predominantly within the striatum. This finding aligns with prior evidence identifying the striatum as a key treatment response biomarker, with robust reductions in cue-evoked activity consistently observed following intervention, particularly in actively treated individuals relative to placebo controls [5, 37]. In addition to the striatum, we observed effects in other subcortical regions, including the amygdala, VTA, and PCC, which collectively support reward/emotion processing, craving, cognitive control, and interoceptive awareness processes central to addiction and recovery [11]. Consistent with our observations, FDCR-based meta-analyses have reported treatment-related reductions in cue reactivity across these regions that may reflect normalization of reward circuitry [37]. Supporting evidence from pharmacological studies further demonstrates that naltrexone reduces alcohol cue–elicited activation in the ventral striatum and heavy drinking [38], clozapine is associated with greater reductions in craving and amygdala reactivity in cannabis use disorder [39], and baclofen attenuates cocaine cue–related activation across limbic and reward-related regions in cocaine-dependent individuals [40]. Together, these findings indicate that reductions in limbic and striatal cue reactivity are sensitive to therapeutic engagement and likely reflect treatment-related modulation of reward and motivational circuitry.

Despite clear evidence of neural target engagement, the absence of a significant time × group interaction in craving ratings may reflect suboptimal stimulation of brain regions most directly involved in regulating drug-related behavior. To enhance behavioral outcomes of dual-site tACS in addiction, future studies could adopt group-level fMRI-informed targeting strategies based on fMRI cue reactivity data. In our previous study on individuals with methamphetamine use disorder (MUD), the commonly used F4–P4 montage did not align with cue-induced frontoparietal connectivity. Instead, the parietal region functionally connected to F4 was closer to Cp4, and F4–Cp4 PPI connectivity correlated with cue-induced craving [33]. This suggests that optimizing stimulation targets based on baseline group-level functional maps may improve the efficacy of dual-site tACS. Additionally, targeting can be individualized. Stimulation sites based on each participant’s cue-reactivity–related functional connectivity may yield better outcomes, as tACS is more effective in regions that are already functionally active or connected [41]. Beyond targeting, stimulation frequency could also be personalized using EEG-based individualized theta frequency to further enhance efficacy [42].

Given the single-session design of this pre-registered clinical trial, our primary focus was on neural target engagement rather than behavioral outcomes such as craving reduction. Based on our recent systematic review, a substantial proportion of tES studies in the field of addiction published up to 2024 (40 out of 96) employed single-session protocols [19]. While several single-session studies have reported significant neural or behavioral effects (e.g., [43-46]), others have reported null findings (e.g., [47-49]), highlighting variability in responsiveness to brief stimulation. Although multi-session or accelerated stimulation protocols may yield more robust behavioral effects [50], the observed neural changes in the present study are consistent with proposed neuroplastic mechanisms underlying tACS. Specifically, offline effects are thought to arise from long-term synaptic plasticity processes—long-term potentiation (LTP) and long-term depression (LTD)—which are driven by spike-timing-dependent plasticity [51]. These mechanisms modulate neural synchronization by strengthening or weakening synaptic connections depending on the relative timing of pre- and post-synaptic activity, and have been shown to support sustained neural effects lasting up to 70 minutes following a single 20-minute tACS session [52-56].

While secondary analyses examined changes in self-reported craving, the primary endpoint of the study was stimulation effects on network-level functional connectivity. As we hypothesized, the theta frequency frontoparietal HD-tACS intervention engaged the targeted network and enhanced synchrony within the right frontoparietal network and across contralateral regions. Notably, increased right frontoparietal PPI connectivity was linked to reduced cue-induced craving, highlighting the network’s role in regulating drug-related behavior. This observation aligns with previous research linking frontoparietal connectivity to both clinical and behavioral outcomes in SUDs. For instance, one study investigating the neural effects of 20 sessions of rTMS over the left DLPFC in individuals with MUD found that, compared to the sham group, those receiving active stimulation exhibited reduced craving and increased functional connectivity between the left DLPFC and the left IPC [57]. Additionally, our findings showed both ipsilateral and contralateral hemispheric engagement, i.e., engagement of left frontal and left parietal nodes with right parietal stimulation site. This interhemispheric interaction may enhance task-related processing in the contralateral cortex through interhemispheric pathways, potentially facilitating executive function [58]. For example, evidence suggests that fewer days since the last substance use (ranging from 1 to 3 days before the scan) were associated with reduced interhemispheric connectivity within the frontoparietal network, suggesting that heightened coupling between these regions may enhance the capacity to reduce craving and sustain abstinence [59].

The Competing Neurobehavioral Decision Systems (CNDS) model proposes that addiction arises from an imbalance between (1) hyperactive reward circuits, such as the medial prefrontal cortex, ventral striatum, VTA, and amygdala, and (2) hypoactive executive control networks, including DLPFC, PPC, and dorsal striatum [60, 61]. Consistent with this model, our study also demonstrated that frontoparietal HD-tACS reduced striatal activity and decreased PPI connectivity between the stimulation sites and both the ventral striatum and amygdala in response to drug cues. These findings support our hypothesis that enhancing frontoparietal synchronization can modulate subcortical cue reactivity in striatal-amygdala circuits.

Frontoparietal tACS successfully engaged brain regions that align with prior neuroimaging research in addiction, which has consistently shown that a disrupted balance between executive control and reward systems contributes to drug-seeking behaviors and relapse risk. For example, resting-state fMRI studies have reported reduced intra-network connectivity within the left frontoparietal network in smokers compared to non-smokers [62], alongside increased connectivity between the frontoparietal and a fronto-striatal network [63]. Similarly, research in alcohol use disorder (AUD) populations using independent component analysis (ICA) found elevated connectivity between the frontoparietal, the salience network, and a subcortical network involving the amygdala and striatum [64]. These patterns highlight the relevance of frontoparietal and subcortical network interactions in addiction and reinforce growing interest in targeting these circuits therapeutically to increase striatal dopamine release [65], suppress reward system activity [66-68], and enhance inhibitory control [69], ultimately contributing to reductions in craving [70].

Our individualized electric field modeling revealed reliable and focal stimulation across participants. Despite inter-individual variability in peak EF coordinates, the use of individualized 10 mm spherical ROIs allowed us to capture subject-specific field distributions while maintaining consistency in our region-of-interest analysis. Importantly, electric field strength within the right frontal stimulation site was significantly correlated with stimulation-induced increases in frontoparietal PPI connectivity in the active group, but not in the sham group. This finding provides preliminary evidence for a dose-dependent neural response to tACS, aligning with prior studies suggesting that higher electric field magnitudes are more likely to induce measurable physiological changes [71, 72]. The absence of this relationship in the sham group supports the specificity of the effect to active stimulation and reinforces the utility of individualized electric field modeling for linking stimulation dose to neural outcomes.

## Limitations and Future Directions

This study has several limitations that warrant consideration. (1) Participants were recruited from a residential treatment program serving both male and female individuals; however, the number of female cases in the center was very low due to structural and geographical reasons, so we ended up recruiting a male-only group. This recruitment reduced the heterogeneity of the collected data, given evidence for sex-dependent responses to brain stimulation [73, 74] and addiction-related neural processes [75]. However, this made a significant limitation to the study, and the generalizability of its findings to female populations remains to be determined. Future studies should aim to include sex-balanced or mixed-gender cohorts, older adults with SUD, individuals outside of residential treatment, or those with differing abstinence durations, and ideally replicate these findings in an independent sample that includes female participants to assess sex-specific effects.

(2) Although we observed significant neural effects, the use of a single 20-minute tACS session may have been insufficient to produce robust behavioral changes. In future studies, multi-session interventions may be necessary to achieve durable changes in craving or drug use behavior, which would also strengthen the clinical relevance of tACS-based treatments. Moreover, while cue-induced craving is a widely used proxy for relapse risk, it remains a subjective measure. Future trials should include long-term follow-up assessments to examine whether neural changes translate into improved cognitive control, changes in physiological stress markers, and finally, reductions in relapse rates confirmed with objective measures such as urine toxicology.

(3) In the context of neural response, two acquisition factors that could contribute to variability should be explicitly acknowledged: (I) Data were acquired on two different MRI scanners within LIBR imaging center; however, (1) type and model were identical across scanners, with assignment determined by availability during recruitment; (2) identical acquisition protocols and scanner setups were used, with scans conducted by the same operating team; and (3) all MRI data underwent standard quality control procedures to identify potential scanner-related artifacts. (II) task-based fMRI was smoothed at 4 mm FWHM, whereas resting-state and gPPI connectivity were smoothed at 8 mm FWHM, as these two values are more common in the CONN toolbox for task and rest analyses.

(4) Additionally, PPI analysis is a widely used functional connectivity approach that assesses whether the statistical relationship between a seed region and other brain regions varies as a function of psychological context (i.e., task)[76, 77]. PPI therefore characterizes context-dependent functional coupling or co-activation between regions, rather than directional or causal influences. As emphasized in prior methodological work, PPI does not permit inference about the direction of information flow or whether one region drives activity in another [76, 77]. In contrast, questions regarding causal or directional interactions—such as top-down or bottom-up causal regulatory influences—require alternative approaches, including lesion-based studies[78], concurrent noninvasive brain stimulation and fMRI[79], or model-based effective connectivity methods such as Dynamic Causal Modeling (DCM) or Structural Equation Modeling (SEM)[80]. Future studies that combine neuromodulation with causal inference methods may provide deeper insight into directional mechanisms.

(5) Moreover, although we employed a pre-validated fMRI drug cue-reactivity task [34] that has been successfully used in our prior studies (e.g., [33, 81-85]) and is publicly available as a standardized resource in addiction research (https://github.com/rkuplicki/LIBR_MOCD), the task design included a yellow border that appeared intermittently during each block. Participants were instructed to push any button on the response box when they saw the yellow box to ensure the participant alertness during the exposure to the neutral and drug cues. Importantly, this feature was presented during both drug cue and neutral blocks and was therefore not specific to cue exposure. While the border was not intended as a cognitive probe, its presence may have introduced additional attentional engagement during task performance. While future studies may like to consider further disentangle cue-reactivity–specific neural responses from attentional processes by using fully passive cue-exposure paradigms, we believe this attentional engagement is a part of the ecological validity of the cue exposure and does not interfere with the experimental paradigm of cue reactivity.

(6) In the present study, we employed a pragmatic, one-size-fits-all stimulation approach based on standard EEG coordinates. While this strategy facilitates reproducibility across participants, it does not account for inter-individual variability in functional network organization. Multiple aspects of brain stimulation, including task context, timing (e.g., stimulation duration, frequency and phase offset between frontal and parietal sites), intensity, and target location, may benefit from individual-level optimization [86]. Future studies leveraging individualized neuroimaging-guided targeting (e.g., structural MRI, fMRI, or precision functional mapping) approaches may further enhance network-specific modulation and stimulation efficacy.

(7) Finally, the absence of long-term follow-up limits our understanding of the durability and clinical relevance of the observed neural changes. Incorporating longitudinal designs with behavioral and physiological endpoints would allow for a more comprehensive assessment of the long-term impact of frontoparietal tACS on substance use recovery.

## Conclusion

In this pre-registered, randomized, triple-blind, sham-controlled clinical trial (NCT03907644), we conducted the first dual-site HD-tACS-fMRI study in addiction to evaluate the effects of 6 Hz in-phase frontoparietal stimulation in individuals with OUD. Our primary outcome showed that active stimulation reduced drug cue–induced striatal reactivity, supporting the role of frontoparietal theta-band tACS in modulating cortico-subcortical circuits. Secondary outcome analyses comparing the active and sham groups revealed distinct patterns of task-dependent connectivity following stimulation. Specifically, frontoparietal task-based connectivity showed a higher average post–pre gPPI beta value in the active group compared with sham, indicating a relative strengthening of task-dependent coupling within the frontoparietal network. In contrast, for cortico-subcortical connections involving the ventral striatum and medial amygdala, the average post–pre gPPI beta values were lower in the active group than in the sham group. Importantly, these reductions reflect a relative shift toward more negative gPPI values, consistent with a strengthening of anti-correlated (negative) task-dependent coupling. Although group differences in subjective craving were not significant, craving changes correlated with frontoparietal connectivity only in the active group, suggesting that longer or repeated stimulation may be needed to elicit behavioral effects. Together, these results demonstrate that network-targeted HD-tACS can modulate neural systems involved in craving regulation. Future studies should incorporate multi-session protocols, long-term behavioral and physiological assessments, and functional imaging–guided targeting to evaluate clinical efficacy and support personalized neuromodulation strategies for substance use disorders.

## Data Availability

Structural and functional MRI data generated in this study are available from the corresponding author upon reasonable request. All experimental code and questionnaires are publicly available at: https://github.com/SoleimaniGhazaleh/Frontoparietal-tACS-OUD/tree/main

## Acknowledgement

Authors would like to thank Yoon-Hee Cha for her scientific support, the 12& 12 addiction recovery center staff and peer group counsellors with Brad Collins leadership, and also the LIBR assessment team with Tim Collins leadership and administrative support from the LIBR executive team with Colleen MacCallum leadership. This work was not possible without their dedicated work for innovations in the development of new therapeutics for mental health disorders.

## References

1. Strang, J., et al., Opioid use disorder. Nature reviews Disease primers, 2020. 6(1): p. 3.

2. Paulozzi, L.J., Prescription drug overdoses: a review. Journal of safety research, 2012. 43(4): p. 283–289.

3. Bahji, A., et al., Mortality among people with opioid use disorder: a systematic review and meta-analysis. Journal of addiction medicine, 2020. 14(4): p. e118–e132.

4. Sangchooli, A., et al., Parameter space and potential for biomarker development in 25 years of fMRI drug cue reactivity: a systematic review. JAMA psychiatry, 2024. 81(4): p. 414–425.

5. Vafaie, N. and H. Kober, Association of drug cues and craving with drug use and relapse: a systematic review and meta-analysis. JAMA psychiatry, 2022. 79(7): p. 641–650.

6. Shi, Z., et al., The role of withdrawal in mesocorticolimbic drug cue reactivity in opioid use disorder. Addiction Biology, 2021. 26(4): p. e12977.

7. Zilverstand, A., et al., Neuroimaging impaired response inhibition and salience attribution in human drug addiction: a systematic review. Neuron, 2018. 98(5): p. 886–903.

8. Luijten, M., et al., Disruption of reward processing in addiction: an image-based meta-analysis of functional magnetic resonance imaging studies. JAMA psychiatry, 2017. 74(4): p. 387–398.

9. Goldstein, R.Z. and N.D. Volkow, Drug addiction and its underlying neurobiological basis: neuroimaging evidence for the involvement of the frontal cortex. American journal of Psychiatry, 2002. 159(10): p. 1642–1652.

10. Everitt, B.J. and T.W. Robbins, Neural systems of reinforcement for drug addiction: from actions to habits to compulsion. Nature neuroscience, 2005. 8(11): p. 1481–1489.

11. Hill-Bowen, L.D., et al., The cue-reactivity paradigm: An ensemble of networks driving attention and cognition when viewing drug and natural reward-related stimuli. Neuroscience & Biobehavioral Reviews, 2021. 130: p. 201–213.

12. Pollard, A.A., et al., Functional neuroanatomy of craving in heroin use disorder: voxel-based meta-analysis of functional magnetic resonance imaging (fMRI) drug cue reactivity studies. The American Journal of Drug and Alcohol Abuse, 2023. 49(4): p. 418–430.

13. Naqvi, N.H., et al., Neural correlates of drinking reduction during a clinical trial of cognitive behavioral therapy for alcohol use disorder. Alcohol: Clinical and Experimental Research, 2024. 48(2): p. 260–272.

14. Westbrook, C., et al., Mindful attention reduces neural and self-reported cue-induced craving in smokers. Social cognitive and affective neuroscience, 2013. 8(1): p. 73–84.

15. Franklin, T., et al., Effects of varenicline on smoking cue–triggered neural and craving responses. Archives of general psychiatry, 2011. 68(5): p. 516–526.

16. Ekhtiari, H., et al., Transcranial electrical and magnetic stimulation (tES and TMS) for addiction medicine: A consensus paper on the present state of the science and the road ahead. Neuroscience & Biobehavioral Reviews, 2019.

17. Mehta, D.D., et al., A systematic review and meta-analysis of neuromodulation therapies for substance use disorders. Neuropsychopharmacology, 2023: p. 1–32.

18. Dinur-Klein, L., et al., Smoking cessation induced by deep repetitive transcranial magnetic stimulation of the prefrontal and insular cortices: a prospective, randomized controlled trial. Biological psychiatry, 2014. 76(9): p. 742–749.

19. Soleimani, G., et al., Effectiveness of Noninvasive Brain Stimulation Protocols on Drug Craving and Consumption/Relapse in Substance Use Disorders: A Systematic Review and Meta-analysis of 208 Clinical Trials and 36 Protocols. medRxiv, 2025: p. 2025.09. 21.25335559.

20. Borrione, L., et al., Home-use transcranial direct current stimulation for the treatment of a major depressive episode: a randomized clinical trial. Jama psychiatry, 2024. 81(4): p. 329–337.

21. Fries, P., Rhythms for cognition: communication through coherence. Neuron, 2015. 88(1): p. 220–235.

22. Parkin, B.L., et al., Dynamic network mechanisms of relational integration. Journal of Neuroscience, 2015. 35(20): p. 7660–7673.

23. Helfrich, R.F., et al., Selective modulation of interhemispheric functional connectivity by HD-tACS shapes perception. PLoS biology, 2014. 12(12): p. e1002031.

24. Polanía, R., et al., The importance of timing in segregated theta phase-coupling for cognitive performance. Current Biology, 2012. 22(14): p. 1314–1318.

25. Violante, I.R., et al., Externally induced frontoparietal synchronization modulates network dynamics and enhances working memory performance. elife, 2017. 6: p. e22001.

26. Van Schouwenburg, M.R., et al., No differential effects of two different alpha-band electrical stimulation protocols over fronto-parietal regions on spatial attention. Frontiers in neuroscience, 2018. 12: p. 433.

27. Kleinert, M.-L., C. Szymanski, and V. Müller, Frequency-unspecific effects of θ-tACS related to a visuospatial working memory task. Frontiers in human neuroscience, 2017. 11: p. 367.

28. Biel, A.L., et al., Modulating verbal working memory with fronto-parietal transcranial electric stimulation at theta frequency: Does it work? European Journal of Neuroscience, 2022. 55(2): p. 405–425.

29. Tzvi, E., et al., Classification of EEG signals reveals a focal aftereffect of 10 Hz motor cortex transcranial alternating current stimulation. Cerebral Cortex Communications, 2022. 3(1): p. tgab067.

30. Fehér, K.D., M. Nakataki, and Y. Morishima, Phase-dependent modulation of signal transmission in cortical networks through tACS-induced neural oscillations. Frontiers in human neuroscience, 2017. 11: p. 471.

31. Network, A.A.C.-R.I.A., et al., Parameter Space and Potential for Biomarker Development in 25 Years of fMRI Drug Cue Reactivity: A Systematic Review. 2024.

32. Koob, G.F. and N.D. Volkow, Neurocircuitry of addiction. Neuropsychopharmacology, 2010. 35(1): p. 217–238.

33. Soleimani, G., et al., How structural and functional MRI can inform dual-site tACS parameters: A case study in a clinical population and its pragmatic implications. Brain stimulation, 2022. 15(2): p. 337–351.

34. Ekhtiari, H., et al., Methamphetamine and Opioid Cue Database (MOCD): development and validation. Drug and alcohol dependence, 2020. 209: p. 107941.

35. Franken, I.H., V.M. Hendriks, and W. van den Brink, Initial validation of two opiate craving questionnaires: the Obsessive Compulsive Drug Use Scale and the Desires for Drug Questionnaire. Addictive behaviors, 2002. 27(5): p. 675–685.

36. Lupi, M., et al., Transcranial direct current stimulation in substance use disorders: a systematic review of scientific literature. The Journal of ECT, 2017. 33(3): p. 203–209.

37. Evohr, B., et al., Cue-Elicited Brain Activity and Treatment Outcomes in Substance Use Disorders: A Meta-Analysis. JAMA Network Open, 2025. 8(12): p. e2548809–e2548809.

38. Schacht, J.P., et al., Predictors of naltrexone response in a randomized trial: reward-related brain activation, OPRM1 genotype, and smoking status. Neuropsychopharmacology, 2017. 42(13): p. 2640–2653.

39. Machielsen, M.W., et al., Comparing the effect of clozapine and risperidone on cue reactivity in male patients with schizophrenia and a cannabis use disorder: A randomized fMRI study. Schizophrenia Research, 2018. 194: p. 32–38.

40. Young, K.A., et al., Nipping cue reactivity in the bud: baclofen prevents limbic activation elicited by subliminal drug cues. Journal of Neuroscience, 2014. 34(14): p. 5038–5043.

41. Bikson, M. and A. Rahman, Origins of specificity during tDCS: anatomical, activity-selective, and input-bias mechanisms. Frontiers in human neuroscience, 2013. 7: p. 688.

42. Zhang, D.-W., A. Moraidis, and T. Klingberg, Individually tuned theta HD-tACS improves spatial performance. Brain Stimulation, 2022. 15(6): p. 1439–1447.

43. Boggio, P.S., et al., Prefrontal cortex modulation using transcranial DC stimulation reduces alcohol craving: a double-blind, sham-controlled study. Drug and alcohol dependence, 2008. 92(1-3): p. 55–60.

44. Cai, B., et al., Differential effects of high-definition transcranial direct current stimulation (HD-tDCS) on attentional guidance by working memory in males with substance use disorder according to memory modality. Brain and Cognition, 2024. 177: p. 106149.

45. Lu, J., et al., Modulation of smoker brain activity and functional connectivity by tDCS: A go/no-go task-state fMRI study. Heliyon, 2023. 9(11).

46. Aksu, S., et al., Transcranial direct current stimulation combined with cognitive training improves decision making and executive functions in opioid use disorder: a triple-blind sham-controlled pilot study. Journal of addictive diseases, 2024. 42(2): p. 154–165.

47. Xu, J., et al., Transcranial direct current stimulation reduces negative affect but not cigarette craving in overnight abstinent smokers. Frontiers in psychiatry, 2013. 4: p. 112.

48. Klauss, J., et al., A randomized controlled trial of targeted prefrontal cortex modulation with tDCS in patients with alcohol dependence. International Journal of Neuropsychopharmacology, 2014. 17(11): p. 1793–1803.

49. Smith, R.C., et al., Effects of transcranial direct current stimulation (tDCS) on cognition, symptoms, and smoking in schizophrenia: a randomized controlled study. Schizophrenia research, 2015. 168(1-2): p. 260–266.

50. Song, S., et al., Effects of single-session versus multi-session non-invasive brain stimulation on craving and consumption in individuals with drug addiction, eating disorders or obesity: A meta-analysis. Brain stimulation, 2019. 12(3): p. 606–618.

51. Markram, H., et al., Regulation of synaptic efficacy by coincidence of postsynaptic APs and EPSPs. Science, 1997. 275(5297): p. 213–215.

52. Zaehle, T., S. Rach, and C.S. Herrmann, Transcranial alternating current stimulation enhances individual alpha activity in human EEG. PloS one, 2010. 5(11): p. e13766.

53. Vossen, A., J. Gross, and G. Thut, Alpha power increase after transcranial alternating current stimulation at alpha frequency (α-tACS) reflects plastic changes rather than entrainment. Brain stimulation, 2015. 8(3): p. 499–508.

54. Guerra, A., et al., Boosting the LTP-like plasticity effect of intermittent theta-burst stimulation using gamma transcranial alternating current stimulation. Brain Stimulation, 2018. 11(4): p. 734–742.

55. Wischnewski, M., et al., NMDA receptor-mediated motor cortex plasticity after 20 Hz transcranial alternating current stimulation. Cerebral cortex, 2019. 29(7): p. 2924–2931.

56. Nitsche, M.A. and W. Paulus, Excitability changes induced in the human motor cortex by weak transcranial direct current stimulation. The Journal of physiology, 2000. 527(Pt 3): p. 633.

57. Su, H., et al., Neuroplastic changes in resting-state functional connectivity after rTMS intervention for methamphetamine craving. Neuropharmacology, 2020. 175: p. 108177.

58. Kornfeld, S., et al., Resting-state connectivity and executive functions after pediatric arterial ischemic stroke. NeuroImage: Clinical, 2018. 17: p. 359–367.

59. McCarthy, J.M., et al., Reduced interhemispheric executive control network coupling in men during early cocaine abstinence: a pilot study. Drug and alcohol dependence, 2017. 181: p. 1–4.

60. Hanlon, C.A., et al., What goes up, can come down: Novel brain stimulation paradigms may attenuate craving and craving-related neural circuitry in substance dependent individuals. Brain research, 2015. 1628: p. 199–209.

61. Bickel, W.K., et al., Competing neurobehavioral decision systems theory of cocaine addiction: From mechanisms to therapeutic opportunities. Progress in brain research, 2016. 223: p. 269–293.

62. Weiland, B.J., et al., Reduced executive and default network functional connectivity in cigarette smokers. Human brain mapping, 2015. 36(3): p. 872–882.

63. Janes, A.C., et al., Prefrontal and limbic resting state brain network functional connectivity differs between nicotine-dependent smokers and non-smoking controls. Drug and alcohol dependence, 2012. 125(3): p. 252–259.

64. Zhu, X., et al., Model-free functional connectivity and impulsivity correlates of alcohol dependence: a resting-state study. Addiction biology, 2017. 22(1): p. 206–217.

65. Strafella, A.P., et al., Repetitive transcranial magnetic stimulation of the human prefrontal cortex induces dopamine release in the caudate nucleus. The Journal of neuroscience, 2001. 21(15): p. RC157.

66. Li, X., et al., Acute left prefrontal transcranial magnetic stimulation in depressed patients is associated with immediately increased activity in prefrontal cortical as well as subcortical regions. Biological psychiatry, 2004. 55(9): p. 882–890.

67. Bohning, D., et al., A combined TMS/fMRI study of intensity-dependent TMS over motor cortex. Biological psychiatry, 1999. 45(4): p. 385–394.

68. De Ridder, D., et al., Transient alcohol craving suppression by rTMS of dorsal anterior cingulate: an fMRI and LORETA EEG study. Neuroscience letters, 2011. 496(1): p. 5–10.

69. Feil, J. and A. Zangen, Brain stimulation in the study and treatment of addiction. Neuroscience & Biobehavioral Reviews, 2010. 34(4): p. 559–574.

70. Perkins, K.A., et al., Placebo effects of tobacco smoking and other nicotine intake. Nicotine & Tobacco Research, 2003. 5(5): p. 695–709.

71. Soleimani, G., et al., Dose-Response in Modulating Brain Function with Transcranial Direct Current Stimulation: From Local to Network Levels. medRxiv, 2022: p. 2022.11. 08.22282088.

72. Esmaeilpour, Z., et al., Methodology for tDCS integration with fMRI. Human Brain Mapping, 2019.

73. Brady, K.T. and C.L. Randall, Gender differences in substance use disorders. Psychiatric Clinics of North America, 1999. 22(2): p. 241–252.

74. Rudroff, T., et al., Response variability in transcranial direct current stimulation: why sex matters. Frontiers in Psychiatry, 2020. 11: p. 585.

75. Huang, Y., et al., Sex and hormonal effects on drug cue-reactivity and its regulation in human addiction. Biological Psychiatry, 2025.

76. O’Reilly, J.X., et al., Tools of the trade: psychophysiological interactions and functional connectivity. Social cognitive and affective neuroscience, 2012. 7(5): p. 604–609.

77. Friston, K.J., Functional and effective connectivity: a review. Brain connectivity, 2011. 1(1): p. 13–36.

78. Siddiqi, S.H., et al., Causal mapping of human brain function. Nature reviews neuroscience, 2022. 23(6): p. 361–375.

79. Jiang, J., The causal neuromodulation mechanisms of the left dorsolateral prefrontal cortex on the amygdala. Brain Stimulation: Basic, Translational, and Clinical Research in Neuromodulation, 2023. 16(1): p. 391.

80. Stephan, K.E., et al., Ten simple rules for dynamic causal modeling. Neuroimage, 2010. 49(4): p. 3099–3109.

81. Ekhtiari, H., et al., Transcranial direct current stimulation to modulate fMRI drug cue reactivity in methamphetamine users: a randomized clinical trial. Human Brain Mapping, 2022. 43(17): p. 5340–5357.

82. Soleimani, G., et al., Converging evidence for frontopolar cortex as a target for neuromodulation in addiction treatment. American Journal of Psychiatry, 2024. 181(2): p. 100–114.

83. Soleimani, G., et al., Dose-response in modulating brain function with transcranial direct current stimulation: from local to network levels. PLoS Computational Biology, 2023. 19(10): p. e1011572.

84. Soleimani, G., et al., Targeting VMPFC-amygdala circuit with TMS in substance use disorder: A mechanistic framework. Addiction Biology, 2025. 30(1): p. e70011.

85. Ekhtiari, H., et al., It is never as good the second time around: Brain areas involved in salience processing habituate during repeated drug cue exposure in treatment engaged abstinent methamphetamine and opioid users. Neuroimage, 2021. 238: p. 118180.

86. Soleimani, G., et al., Four dimensions of individualization in brain stimulation for psychiatric disorders: context, target, dose, and timing. Neuropsychopharmacology, 2025: p. 1–14.

