## Supplementary Materials for "Targeting Cortico-Striatal-Amygdalar Networks via Theta-Band Frontoparietal Synchronization in Opioid Use Disorder: A Randomized tACS-fMRI Trial"

CONSORT

TRANSPARENT REPORTING of TRIALS

### CONSORT 2010 Flow Diagram

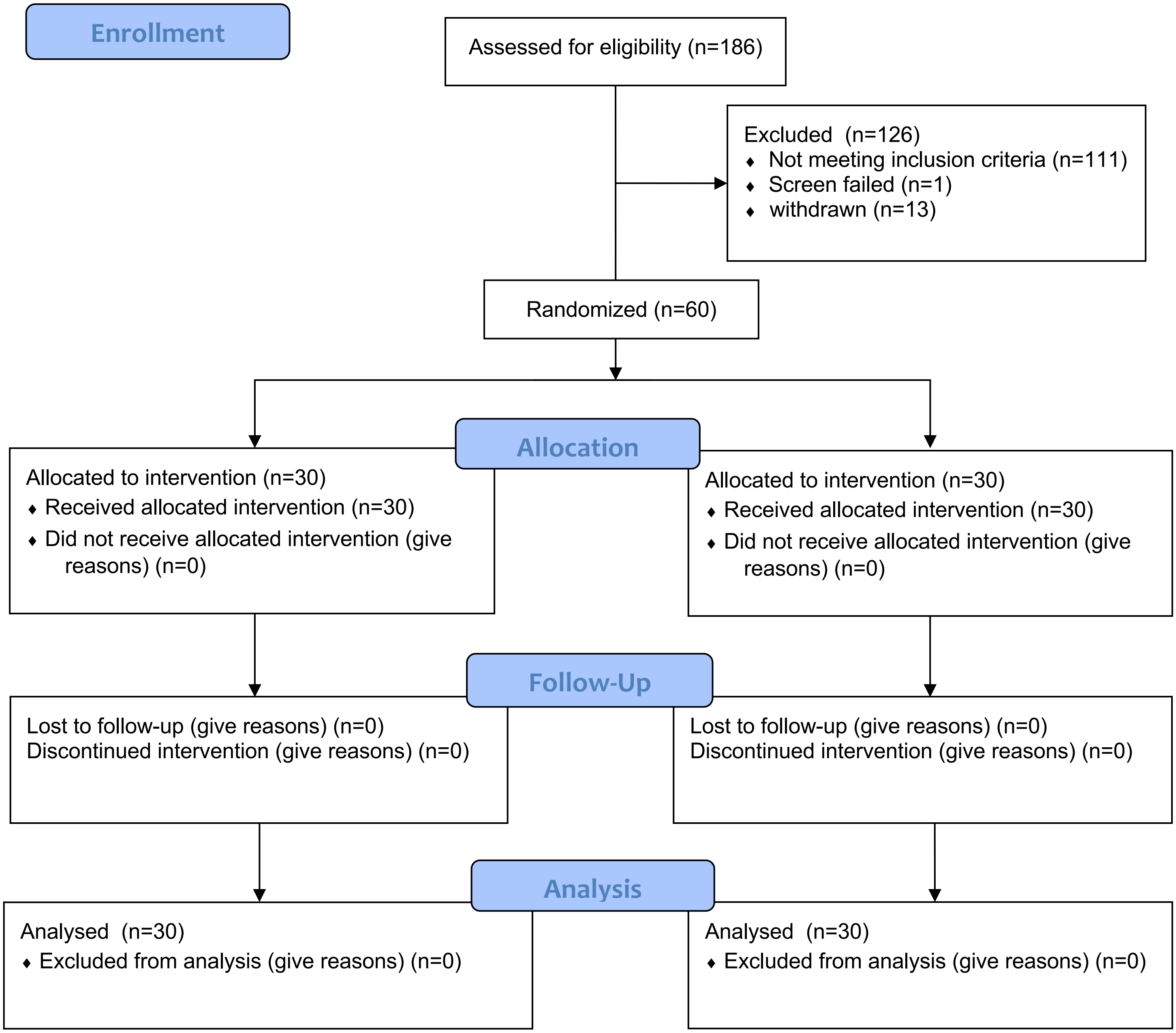

**S1. Inclusion/Exclusion criteria**

To participate in the study, individuals needed to meet several criteria: they had to speak English, have been diagnosed with opioid use disorder (OUD) based on DSM V criteria [1] within the past year using MINI structured interview [2], be enrolled in a residential treatment program for OUD, have abstained from opioids for at least one week, and be willing and able to provide informed consent. On the other hand, certain conditions would disqualify potential participants. These included an unwillingness or inability to complete key parts of the study (such as MRI scans, drug cue ratings, or behavioral assessments), having been opioid-free for more than six months (self-reported), having schizophrenia or bipolar disorder (as identified through the MINI interview), exhibiting active suicidal thoughts or plans (determined through self-report or assessment by the study team), and testing positive for amphetamines, opioids, cannabis, alcohol, phencyclidine, or cocaine (confirmed by breath analyzer and urine tests).

**S2. More details on experiment design**

Neuroimaging data, including structural MRI (T1-weighted MRI), resting-state fMRI, and task-based fMRI with a block-designed task, were collected using a pre-stimulation/post-stimulation design. Subjective craving for opioid use was measured using a Visual Analog Scale (VAS) scored from 0 to 100 at six different time points: baseline (t0), immediately before (t1 and t3), immediately after (t2 and t4) each fMRI scan, and the day after data collection (t5). Adverse effects of tACS were tracked by asking participants to complete a questionnaire both at the end of the scanning session and the following day. This questionnaire used a Likert scale from 0 to 5 (0 = none, 1 = very mild, 2 = mild, 3 = moderate, 4 = severe, 5 = very severe) to rate 10 different symptoms. The desire for Drug Questionnaire (DDQ) [3] with three main sub-scores, that is, desire and intention, negative, and control, was collected at four different time points (t0, t1, t4, and t5). Additionally, to assess the effectiveness of blinding, participants were asked at the end of the stimulation session whether they believed they had received sham or active stimulation and how confident they were in their assessment.

**S3. More details on tACS parameters and Randomization**

A 32-channel StarStim device was used to apply transcranial alternating current stimulation (tACS). All high-definition electrodes were silver/silver chloride and were wetted with conductive gel prior to placement. Surface landmarks and EEG caps were used to position the electrodes on the scalp. Electrodes were secured using head caps with predefined openings corresponding to the electrode locations.

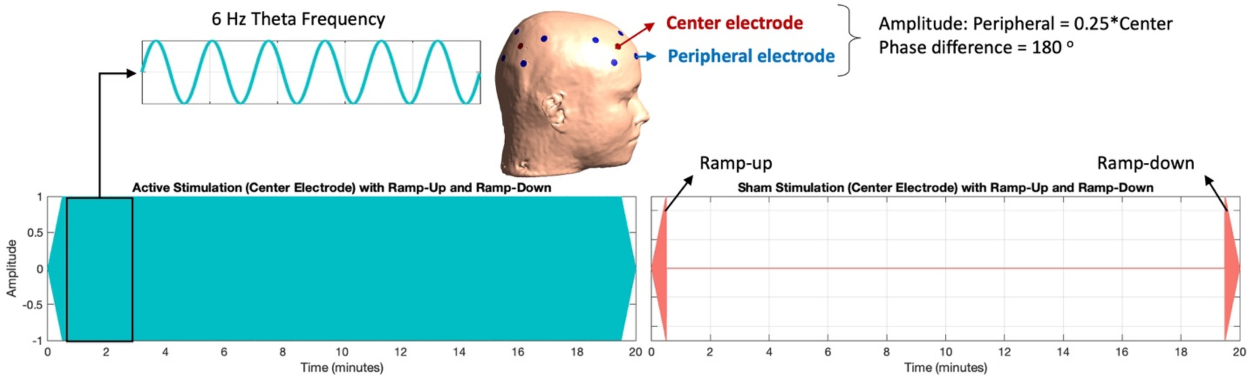

**Figure S1. Active vs Sham stimulation waveform.** Active stimulation is 20 minutes of in-phase frontoparietal tACS at a theta frequency (6 Hz) with 2 mA peak-to-peak and 30 seconds ramp up and ramp down over the right center electrodes (F4 and P4). The four peripheral electrodes at each site received 0.5 mA peak-to-peak with a 180-degree phase difference from the center electrode. During sham stimulation, the device only provides ramp-up and ramp-down stimulation.

To maintain the triple-blind nature of the study—such that participants, the experimenter administering stimulation, and data analysts remained unaware of the stimulation condition—the stimulation device was operated in built-in study mode, in which active and sham protocols were encoded using device-generated numeric codes and stimulation parameters were not modifiable by the experimenter. Stimulation programs were assigned by a study manager who was not involved in data collection or analysis and who alone had access to the code key. During stimulation, the experimenter interface displayed only these non-informative labels and numerical identifiers, with no visual or textual indicators of stimulation type. These codes corresponded to pre-coded stimulation waveforms that were uploaded to the device, ensuring that neither the participant nor the experimenter could infer the stimulation condition at any point during the experiment.

### S4. More details on MRI parameters

Resting-state fMRI data were collected in an 8-minute run both before and after the stimulation sessions, preceding the task-based fMRI. During the resting-state scans, participants were instructed to keep their eyes fixed on a cross displayed on the screen and to try to clear their minds of any specific thoughts. Following the resting-state fMRI, participants performed a standard drug vs. neutral cue reactivity task, which employed a block design with two sets of distinct but equivalent pictures (opioid and neutral as validated here [4]). The task lasted approximately 6.5 minutes and consisted of four blocks of neutral pictures and four blocks of opioid-related pictures. Each block contained six pictures from the same category (either opioid or neutral), with each picture shown for 5 seconds, separated by a 0.2-second blank interstimulus interval. At the end of each block, participants rated their craving levels. A visual fixation point was presented between two consecutive blocks for 8–12 seconds.

Structural and functional MRI data were collected using two identical GE MRI 750 3T scanners. The structural MRI parameters included a TR/TE of 5/2.012 ms, a field of view (FOV) of 24 x 192 mm per slice, and a 256x256 matrix, producing voxel dimensions of 0.938 x 0.9 mm with 186 axial slices for T1-weighted images. These T1-weighted images were utilized to generate computational head models for each participant. The parameters for resting state run were a TR/TE of 2000/27 ms, an FOV of 240 mm per slice, and a 128x128 matrix, yielding voxel dimensions of 1.857x1.857x2.9 mm, with 39 axial slices and 240 repetitions. For task-based fMRI, the parameters were set to a TR/TE of 2000/27 ms, an FOV of 240 mm per slice, and a 128x128 matrix, resulting in voxel dimensions of 1.857x1.857x2.9 mm, with 39 axial slices and 196 repetitions.

### S5. fMRI Drug Cue Reactivity (FDCR) task design

The opioid cue reactivity task was administered following each resting-state scan, maintaining the same parameters, except it included 196 repetitions. This task involved exposing participants to opioid-related and neutral images in a block design, using distinct but equivalent pictures validated by Ekhtiari, Kuplicki, Pruthi, and Paulus (2020).

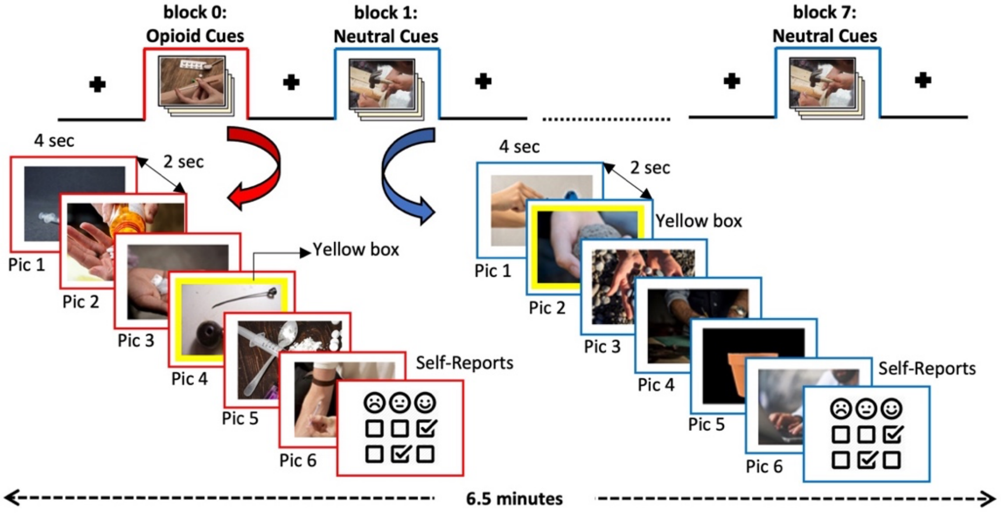


**Figure S2. Schematic of the fMRI Drug Cue Reactivity Task Paradigm.** This diagram illustrates the block design used in the fMRI task, featuring examples of neutral stimuli and opioid cues. The task lasted around 6.5 minutes, comprising 4 blocks of neutral images and 4 blocks of opioid images.

The task lasted approximately 6.5 minutes and comprised four blocks of opioid images and four blocks of neutral images. Each block featured six pictures from the same category (opioid or neutral), each displayed for 5 seconds with a 0.2-second blank interval between images. The opioid cue images included visual representations of heroin and other opioids and their paraphernalia. Images in each category (opioid or neutral) were subdivided into six content-based categories: (i) drug or control neutral object alone, (ii) drug or control neutral object with hands, (iii) drug related (paraphernalia) and or control neutral tools or instruments, (iv) tools or instruments with hands, (v) tools or instruments with hands in use action, and (vi) drug-related or neutral activities including faces.

Between each block, a visual fixation point was shown for 8 to 12 seconds. After each block, participants rated their current opioid craving level on a scale from 1 (lowest) to 4 (highest). The response times for these ratings were recorded. Additionally, during each block, a yellow box would occasionally appear around one of the six pictures, prompting participants to press a button as quickly as possible, with their reaction times recorded. Given the timing of the resting-state fMRI, and the fact that MRI data were collected immediately before and after the stimulation session, the second task-based fMRI was conducted about 8 minutes after the end of the stimulation. This interval included the time required to move the participant from the scanner to the stimulation room and back, as the stimulation was performed in a room adjacent to the scanner room.

#### S6.1. fMRI Preprocessing

**Functional Activity:** Functional data analysis was conducted using the AFNI software package. Preprocessing steps were applied, including the removal of the first three pre-steady state images. The following procedures were performed: despiking, slice timing correction, realignment, transformation to MNI space, and Gaussian FWHM smoothing with a kernel size of 4 mm. To account for potential confounding factors, three polynomial terms and the six motion parameters were regressed out during the analysis. Additionally, TRs with excessive motion were identified and censored based on a criterion defined as the Euclidean norm of the derivative of the six motion parameters exceeding 0.3.

**Functional Connectivity:** Functional connectivity analysis was performed in the CONN toolbox. Preprocessing the data involved several key steps. First, all scans were realigned and unwarped, co-registering them to the first volume. Next, slice timing correction was applied. Outlier detection was then performed using the Artifact Detection Tools (ART) scrubbing procedure in the CONN toolbox, which identified potential motion outliers based on 12 motion parameters (x, y, z, pitch, roll, yaw, and their first-order derivatives). Scans with framewise displacement over 0.9 mm were flagged, and subjects with more than 25% of their scans flagged were excluded from further analysis. Functional and structural data were subsequently normalized to MNI space, with functional data referenced against the mean BOLD signal and structural data against T1-weighted MRI. These data were segmented into gray matter, white matter, and CSF, and each segmented tissue type was visually inspected to ensure no incoherent deformation. Finally, functional smoothing was performed using an 8 mm full-width half-maximum (FWHM) Gaussian kernel to enhance the signal-to-noise ratio and improve the validity of subsequent analyses.

**S6.2. More Details on Seed Definition**

**A computational head modeling approach:** For each individual, the location of the 99th percentile of the electric field was calculated over the frontal and parietal cortices and transformed into MNI space. Spheres with a radius of 10 mm were placed around each location. These spheres were then combined with the MNI mask to ensure that analyses did not include signals from non-brain or white matter voxels. All of these steps were also replicated for the left hemisphere as well.

**Atlas-based parcellation:** To analyze cortico–subcortical connections, we applied the Brainnetome atlas, a fine-grained, functionally informed parcellation, to the fMRI data. Each Brainnetome subregion was saved for subsequent functional analyses. The Brainnetome atlas was used for cortical and amygdala parcellation because it provides detailed and well-validated functional subdivisions of these regions. For striatal analyses, however, we used the Harvard–Oxford subcortical atlas, as it provides anatomically intuitive and widely used subdivisions of the striatum (e.g., ventral caudate, dorsal caudate, putamen, globus pallidus, and nucleus accumbens), which are more suitable for hypothesis-driven cortico-striatal connectivity analyses. Averaged locations for frontoparietal regions obtained from the highest electric fields, amygdala subregions (Brainnetome parcels), and striatum subregions (Harvard–Oxford parcels) used as the main nodes in our analyses are visualized in Figure S3.

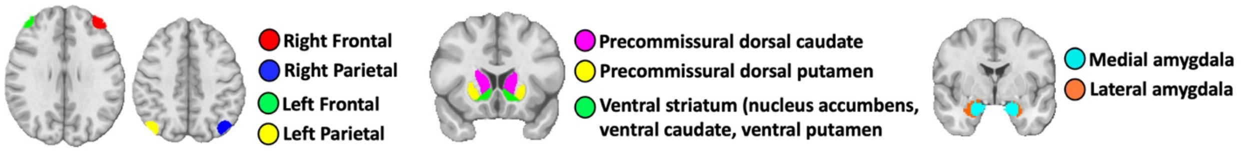

**Figure S3. Main seeds that are used for further fMRI analysis or electric field calculations.** Frontoparietal nodes were obtained by defining 10 mm spheres around the 99$^{th}$ percentile of the EFs for each person, Striatum subregions were obtained from the Oxford-Harvard atlas, and Amygdala subregions were obtained from the Brainnetome atlas parcellation.

#### S6.3. More details on electric field simulations

High-resolution T1-weighted MRI data were used along with SimNIBS 3.2 software to create personalized computational head models. These models included six primary tissue types: white matter (WM), gray matter (GM), cerebrospinal fluid (CSF), skull, eye vitreous bodies, and skin. The segmentation process was performed using the "headreco" function in SimNIBS. Each segmented tissue was visually checked for accuracy against the T1-weighted MR images. Tetrahedral volume meshes, derived from these segmented surfaces, were generated for each model, containing about 5-6 million tetrahedra. For simulation purposes, 2 mm sphere electrode pads with a thickness of 4 mm were modeled on the scalp to represent the electrode locations. Different conductivity values were assigned to each head model based on a distribution representing human tissue properties to account for individual variability in tissue conductivities. Specifically, the conductivities of the scalp, skull, and gray matter were based on the β(3,3) distribution, commonly used in uncertainty modeling. In contrast, specific values were assigned to the conductivities for CSF, white matter, and eyes, as their influence on the resulting electric field is minimal. The resulting conductivity values were as follows (all S/m): Skull: 0.002 to 0.03, Skin: 0.2 to 0.6, CSF: 1.66, Eye: 0.5, Gray Matter: 0.1 to 0.6, and White Matter: 0.14.

### S7. Participants' substance use history

To further characterize participants' substance use profiles, we assessed the duration of use (in years) across various substances and alcohol.

**Table S1. Years of Substance/Alcohol Use**

| Question | Sham (n = 30) Mean ± SD | Active (n = 30) Mean ± SD | P Value |
|---|---|---|---|
| Heroin | 7.73 ± 6.58 (n=26) | 7.59 ± 6.06 (n=27) | 0.937 |
| Opioids/Analgesics | 12.18 ± 7.77 (n=28) | 14.11 ± 7.29 (n=27) | 0.346 |
| Alcohol (any) | 16.26 ± 7.99 (n=30) | 16.30 ± 9.07 (n=27) | 0.297 |
| Alcohol (to intoxication) | 13.48 ± 11.78 (n=29) | 13.77 ± 9.34 (n=26) | 0.921 |
| Multi-Substance | 13.07 ± 8.36 (n=29) | 15.21 ± 9.04 (n=29) | 0.356 |
| Cannabis | 16.07 ± 8.49 (n=29) | 16.04 ± 9.46 (n=27) | 0.989 |
| Amphetamines | 9.56 ± 7.85 (n=25) | 10.81 ± 7.94 (n=26) | 0.575 |
| Cocaine | 5.96 ± 6.44 (n=23) | 6.74 ± 7.99 (n=23) | 0.716 |
| Sedative/Tranquilizer | 9.81 ± 9.09 (n=21) | 6.77 ± 10.85 (n=22) | 0.474 |
| Hallucinogen | 4.74 ± 5.26 (n=17) | 4.58 ± 4.56 (n=19) | 0.948 |
| Barbiturate | 2.00 ± 00 (n=1) | 7.71 ± 8.36 (n=7) | 0.546 |
| Inhalant | 2.00 ± 1.73 (n=3) | 3.29 ± 3.99 (n=7) | 0.615 |

### S8. Blindness and Adverse Effects

Using a structured approach, participants were assessed for blindness and side effects immediately after and on the day following the stimulation. To assess the integrity of blinding, participants were asked after the stimulation session (after-post fMRI, t4) and again the following day (t5) to guess whether they had received real or sham stimulation. They also rated their confidence in this guess on a scale from 0 to 100. In addition, they were asked whether they believed the stimulation reduced their craving (Yes/No) and to rate the perceived effectiveness of the stimulation in reducing craving on a scale from 0 to 100. Table S2 summarizes these responses for both the sham and active stimulation groups. Overall, the majority of participants in both groups guessed they had received real stimulation, with no significant differences in guess accuracy across groups or timepoints. Confidence in guesses was significantly higher in the active group on Day After (t5), but no group differences were observed in the belief that stimulation reduced craving or in the perceived effectiveness ratings.

**Table S2. Blindness efficacy**

| | Question | Sham (n = 30) Mean ± SD | Active (n = 30) Mean ± SD | P Value |
|---|---|---|---|---|
| **Day After (t5)** | Guess of stimulation (Real vs Sham) | 27 Real (90.0%) | 23 Real (82.1%) | 0.464 |
| | Confidence in guess (0-100) | 70.37 ± 24.54 | 83.11 ± 17.48 | 0.028 |
| | Belief stimulation reduced craving (0=No, 1=Yes) | 0.72 ± 0.45 (Yes=23) | 0.71 ± 0.46 (Yes=24) | 0.936 |
| | Perceived effectiveness (0-100) | 59.17 ± 27.43 | 57.32 ± 31.64 | 0.813 |
| **After Post-Stim fMRI (t4)** | Guess of stimulation (Real vs Sham) | 24 Real (77.4%) | 25 Real (86.2%) | 0.509 |
| | Confidence in guess (0-100) | 77.35 ± 18.35 | 76.76 ± 22.28 | 0.910 |
| | Belief stimulation reduced craving (0=No, 1=Yes) | 0.74 ± 0.44 (Yes=21) | 0.83 ± 0.38 (Yes=20) | 0.430 |
| | Perceived effectiveness (0-100) | 62.19 ± 26.10 | 62.55 ± 21.52 | 0.954 |

They were asked to report whether they experienced any of the following symptoms: headache, neck pain, scalp pain, tingling, itching, burning sensation, redness, sleepiness, trouble concentrating, or acute mood changes (responding with "Yes" or "No," e.g., "Do you have a headache? Yes/No"). If they did experience any side effects, they were then asked to rate the severity (e.g., "Please describe the severity: absent/mild/moderate/severe"). Additionally, participants were queried about whether they believed the side effects were related to the tACS treatment, with responses quantified across five levels (e.g., "Is this related to tACS? None/remote/possible/probable/definite"). Results are reported in Table S3.

**Table S3. Adverse effects**

| A: Sham, B: Active | After Post fMRI (t4) | | | | | | | | | | | Day After (t5) | | | | | | | | | | |
|---|---|---|---|---|---|---|---|---|---|---|---|---|---|---|---|---|---|---|---|---|---|---|
| | Side effects | | Severity | | | | Relatedness | | | | | Side effect | | Severity | | | | Relatedness | | | | |
| | No | Yes | Absent | Mild | Moderate | Severe | None | Remote | Possible | Probable | Definite | No | Yes | Absent | Mild | Moderate | Severe | None | Remote | Possible | Probable | Definite |
| **Headache** | | | | | | | | | | | | | | | | | | | | | | |
| A | 25 | 5 | 1 | 3 | 1 | 0 | 1 | 0 | 3 | 0 | 0 | 26 | 4 | 0 | 4 | 0 | 0 | 3 | 0 | 1 | 0 | 0 |
| B | 27 | 3 | 0 | 3 | 0 | 0 | 1 | 0 | 2 | 0 | 0 | 26 | 4 | 1 | 2 | 1 | 0 | 0 | 1 | 1 | 1 | 0 |
| **Neck pain** | | | | | | | | | | | | | | | | | | | | | | |
| A | 28 | 2 | 0 | 2 | 0 | 0 | 0 | 0 | 2 | 0 | 0 | 28 | 2 | 0 | 2 | 0 | 0 | 1 | 0 | 1 | 0 | 0 |
| B | 25 | 5 | 0 | 4 | 1 | 0 | 2 | 0 | 2 | 1 | 0 | 25 | 5 | 0 | 3 | 2 | 0 | 2 | 1 | 1 | 1 | 0 |
| **Scalp pain** | | | | | | | | | | | | | | | | | | | | | | |
| A | 29 | 1 | 0 | 1 | 0 | 0 | 0 | 0 | 0 | 1 | 0 | 29 | 1 | 0 | 1 | 0 | 0 | 0 | 0 | 1 | 0 | 0 |
| B | 29 | 1 | 0 | 1 | 0 | 0 | 0 | 0 | 0 | 0 | 1 | 30 | 0 | 0 | 0 | 0 | 0 | 0 | 0 | 0 | 0 | 0 |
| **Tingling** | | | | | | | | | | | | | | | | | | | | | | |
| A | 26 | 4 | 1 | 3 | 0 | 0 | 1 | 0 | 0 | 2 | 0 | 29 | 1 | 0 | 1 | 0 | 0 | 0 | 0 | 0 | 1 | 0 |
| B | 26 | 4 | 1 | 3 | 0 | 0 | 0 | 0 | 1 | 2 | 0 | 29 | 1 | 0 | 1 | 0 | 0 | 1 | 0 | 0 | 0 | 0 |
| **Itching** | | | | | | | | | | | | | | | | | | | | | | |

| | | | | | | | | | | | | | | | | | | | | | | |
|---|---|---|---|---|---|---|---|---|---|---|---|---|---|---|---|---|---|---|---|---|---|---|
| A | 26 | 4 | 1 | 3 | 0 | 0 | 1 | 0 | 1 | 1 | 0 | 30 | 0 | 0 | 0 | 0 | 0 | 0 | 0 | 0 | 0 | 0 |
| B | 23 | 7 | 1 | 4 | 2 | 0 | 1 | 1 | 3 | 1 | 0 | 29 | 1 | 0 | 1 | 0 | 0 | 1 | 0 | 0 | 0 | 0 |
| **Burning sensation** | | | | | | | | | | | | | | | | | | | | | | |
| A | 28 | 2 | 0 | 1 | 1 | 0 | 0 | 0 | 1 | 0 | 1 | 30 | 0 | 0 | 0 | 0 | 0 | 0 | 0 | 0 | 0 | 0 |
| B | 28 | 2 | 0 | 2 | 0 | 0 | 0 | 0 | 1 | 0 | 1 | 30 | 0 | 0 | 0 | 0 | 0 | 0 | 0 | 0 | 0 | 0 |
| **Skin redness** | | | | | | | | | | | | | | | | | | | | | | |
| A | 30 | 0 | 0 | 0 | 0 | 0 | 0 | 0 | 0 | 0 | 0 | 30 | 0 | 0 | 0 | 0 | 0 | 0 | 0 | 0 | 0 | 0 |
| B | 30 | 0 | 0 | 0 | 0 | 0 | 0 | 0 | 0 | 0 | 0 | 29 | 1 | 0 | 0 | 0 | 1 | 1 | 0 | 0 | 0 | 0 |
| **Sleepiness** | | | | | | | | | | | | | | | | | | | | | | |
| A | 16 | 14 | 1 | 8 | 5 | 0 | 8 | 3 | 1 | 1 | 0 | 27 | 3 | 1 | 2 | 0 | 0 | 2 | 0 | 0 | 0 | 0 |
| B | 17 | 13 | 0 | 12 | 1 | 0 | 4 | 2 | 6 | 0 | 1 | 22 | 8 | 1 | 6 | 1 | 0 | 4 | 1 | 2 | 0 | 0 |
| **Trouble concentrating** | | | | | | | | | | | | | | | | | | | | | | |
| A | 27 | 3 | 0 | 2 | 0 | 1 | 2 | 0 | 0 | 1 | 0 | 29 | 1 | 0 | 0 | 0 | 1 | 1 | 0 | 0 | 0 | 0 |
| B | 30 | 0 | 0 | 0 | 0 | 0 | 0 | 0 | 0 | 0 | 0 | 26 | 4 | 0 | 2 | 2 | 0 | 3 | 0 | 1 | 0 | 0 |
| **Acute mood change** | | | | | | | | | | | | | | | | | | | | | | |
| A | 24 | 6 | 0 | 5 | 1 | 0 | 1 | 1 | 3 | 1 | 0 | 29 | 1 | 0 | 1 | 0 | 0 | 0 | 0 | 0 | 0 | 1 |
| B | 25 | 5 | 0 | 3 | 2 | 0 | 1 | 0 | 2 | 1 | 1 | 26 | 4 | 0 | 2 | 1 | 0 | 1 | 0 | 1 | 1 | 0 |

### S9. More details on behavioral Results

#### S9.1. Craving scores (How much craving do you have right now?)

0 = no craving, 100 = highest craving**:** At baseline, both groups showed similar levels of craving (sham: 32.23 ± 30.64, active: 33.72 ± 28.25; p = 0.84445). Before the pre-fMRI session, cravings slightly decreased in both groups (sham: 29.03 ± 29.13, active: 32.45 ± 27.47; p = 0.6419). Following the pre-fMRI session, cravings increased notably in both groups (sham: 57.10 ± 29.7, active: 54.75 ± 29.35; p = 0.76152). Before the post-fMRI session (t3), craving was slightly higher in the sham (30.61 ± 25.19) compared to the active (22.57 ± 20.92; p = 0.18614). After the post-fMRI session, cravings decreased further in both groups (sham: 27.68 ± 24.09, active: 26.07 ± 29.52; p = 0.82095). By the day after the session, cravings continued to decrease in both groups (sham: 15.63 ± 20.97, active: 18.59 ± 20.85; p = 0.58971).

**Within-group post-task craving change (t4 − t3):** To assess whether craving changed during the post-stimulation FDCR task within each group, post-task craving change (t4 − t3) was tested against zero separately for the sham and active groups. No significant change was observed in both active (mean = −2.94, SD = 26.62; t(30) = −0.61, p = 0.54; sign-rank p = 0.46) and sham (mean = 3.45, SD = 23.04; t(28) = 0.81, p = 0.43; sign-rank p = 0.59) groups. These findings indicate that neither group exhibited a reliable within-group change in craving during the post-stimulation task window.

**Between-group comparison of post-task craving change:** To determine whether post-task craving change differed between groups, post-task craving change (t4 − t3) was compared between the sham and active groups using a Welch two-sample t-test. No significant difference was observed between groups (t(57.67) = −0.99, p = 0.32). This result indicates that post-task craving changes did not differ reliably between sham and active stimulation groups, arguing against a differential placebo-related effect during the post-stimulation task.

**Linear mixed-effects analysis of task-related craving change:** To evaluate task-related craving changes while accounting for within-subject variability, a linear mixed-effects model was fitted with Group (sham vs active), session (pre-stimulation vs post-stimulation), and their interaction as fixed effects, and participant as a random effect. The model revealed a significant main effect of MRI session ($F(1,116) = 17.49$, $p < 0.001$), indicating that craving levels were lower during the post-stimulation MRI compared with the pre-stimulation MRI across both groups. In contrast, neither the main effect of Group ($p = 0.33$) nor the Group × time interaction ($p = 0.20$) reached significance. These findings suggest that the observed reduction in craving from pre- to post reflects a general within-session or task-related effect rather than a stimulation-specific or placebo-driven modulation.

**Follow-up day (t5) group comparisons:** To address the observation that craving appeared descriptively lower in the sham group at follow-up, craving scores at t5 were compared between groups. No significant group difference was observed ($t(57.64) = -0.53$, $p = 0.60$). Furthermore, changes in craving from baseline to follow-up (t5 − t1) did not differ between the sham and active groups ($t(57.38) = 0.08$, $p = 0.94$), with both groups showing comparable reductions in craving relative to baseline. These results indicate that the numerically lower mean craving score in the sham group at follow-up does not reflect a statistically meaningful group difference.

#### S9.2. Control scores (How much control do you have on your craving right now?)

0 = no control, 100 = highest control**:** At baseline, both groups showed similar levels of control (sham: 50.03 ± 34.32, active: 43.72 ± 31.01; $p = 0.4576$). Before the pre-fMRI session, control scores were slightly lower in sham (38.16 ± 33.76) compared to active (43.83 ± 31.77; $p = 0.50565$). Following the pre-fMRI session, control scores slightly increased in both groups (sham: 44.39 ± 26.99, active: 48.54 ± 31.73; $p = 0.59282$). Before the post-fMRI session, control scores showed a slight decrease in sham (41.06 ± 32.89) compared to active (35.00 ± 32.43; $p = 0.47908$). After the post-fMRI session, control scores decreased further in both groups (sham: 33.61 ± 29.86, active: 31.07 ± 33.38; $p = 0.76004$). By the day after the session, control scores continued to decrease in both groups (sham: 22.93 ± 27.24, active: 20.52 ± 28.13; $p = 0.73888$). Similar to the results reported for craving in the main manuscript, for control scores, time had a significant effect ($F(5, 344) = 5.52$, $p = 0.0001$), suggesting changes across assessment points, while the effect of group ($F(1, 344) = 0.142$, $p = 0.7066$) and the interaction between time and group ($F(5, 344) = 0.393$, $p = 0.8535$) were not significant. These findings suggest fluctuating craving and control scores across different assessment points with no significant differences between sham and active groups.

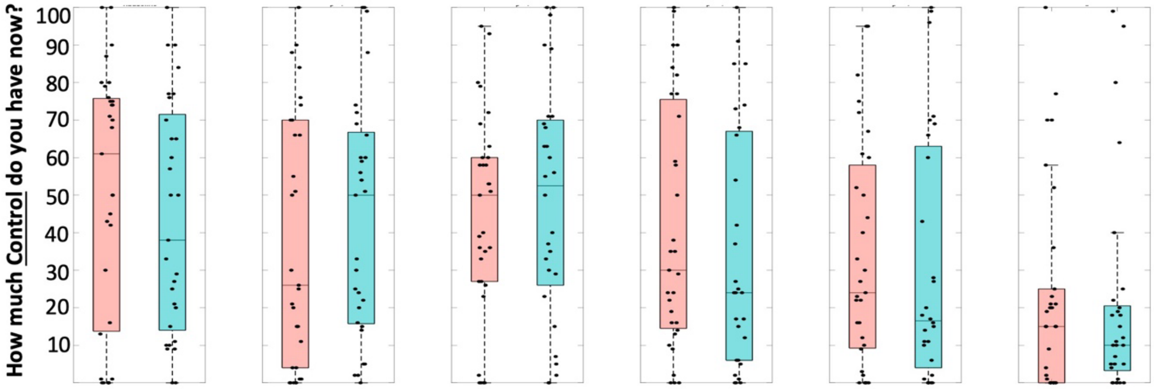


**Figure S4. Control in Craving Assessment and Results.** Based on VAS (0–100), self-reported control over craving was collected at six separate time points and reported for each group. Box plots indicate median values for craving (first row) and control over craving (second row), with dots representing individual data points. Boxes are color-coded for active (blue) and sham (red) groups.

#### S9.3. Desire for Drug Cue Questionnaire (DDQ)

Current drug craving was assessed using the Desire for Drug Questionnaire – Now (DDQ–Now), a 13-item self-report measure designed to capture momentary craving states. Participants rated each item on a 7-point Likert scale ranging from 1 (Strongly Disagree) to 7 (Strongly Agree).

1. Using drugs would be satisfying now.
2. I would consider using drugs now.
3. If I started using drugs now, I would be able to stop.
4. I would do almost anything to use drugs now.
5. I would feel less worried about my daily problems if I used drugs now.
6. My desire to use drugs now seems overwhelming.
7. I could easily limit how much drugs I would use if I used now.
8. I would feel as if all the bad things in my life had disappeared if I used drugs now.
9. I want drugs so much I can almost taste it.
10. Using drugs now would make me feel less tense.
11. Even major problems in my life would not bother me if I used drugs now.
12. Using drugs would be pleasant now.
13. I am going to use drugs as soon as I possibly can.

The questionnaire yields three subscale scores reflecting distinct components of craving.

##### Desire and Intention (DDQ Desire)

The Desire/Intention subscale assesses the intensity of craving and readiness to use drugs (items 1, 2, 4, 6, 9, 12, and 13), capturing motivational drive and behavioral intention. This subscale reflects the motivational drive and conscious intention to use drugs at the present moment. It captures how strongly an individual wants drugs, how compelling the urge feels, and whether they are inclined to act on that urge.

##### Negative Reinforcement (DDQ Negative)

The Negative Reinforcement subscale reflects craving driven by relief from negative emotional states (items 5, 8, 10, and 11), indexing the expectation that drug use would reduce tension, worry, or distress. This subscale represents craving driven by relief from negative emotional or physical states**.** It reflects the expectation that drug use would reduce distress, tension, worry, or life problems.

##### Perceived Control Over Use (DDQ Control)

The Control subscale measures perceived ability to regulate or stop drug use if initiated (items 3 and 7), reflecting perceived self-control over consumption. The control subscale measures perceived self-regulatory capacity over drug use**.** It reflects how much individuals believe they could stop or limit drug use if they began using at the present moment.

Subscale scores were computed as the mean of their respective items and rounded to two decimal places for analysis and reporting. Higher scores on the Desire/Intention and Negative Reinforcement subscales indicate stronger craving and greater affect-driven motivation to use, whereas higher scores on the Control subscale indicate greater perceived ability to limit or stop drug use.

The results of the interaction effect analysis for the DDQ scores are presented in the table. For the "control of craving" score, the Group × Session interaction effect was insignificant (F = 0.01, p = 0.99), indicating no significant difference between the active and sham groups over time. Similarly, the "desire for drug" score showed a non-significant Group × Session interaction effect (F = 2.72, p = 0.10). However, there was a trend toward significance, suggesting a potential decrease in the desire for drugs in the active group compared to the sham group. The "negative affect" score also exhibited a non-significant Group × Session interaction effect (F = 0.64, p = 0.47), indicating no significant change in negative affect scores between the groups over time.

**Table S4. Post hoc analysis for DDQ**

| Session | DDQ Question | Sham | Active | p-value | t-statistic |
|---|---|---|---|---|---|
| baseline | Control score | 1.76 ± 1.30 | 2.14 ± 1.59 | 0.3127 | -1.0185 |
| baseline | Desire score | 2.19 ± 1.11 | 2.00 ± 0.96 | 0.4824 | 0.707 |
| Baseline | Negative score | 2.98 ± 2.01 | 3.03 ± 1.46 | 0.9121 | -0.1109 |
| Before pre fMRI | Control score | 2.31 ± 1.65 | 1.83 ± 1.22 | 0.209 | 1.2704 |
| Before pre fMRI | Desire score | 2.15 ± 1.30 | 1.86 ± 0.87 | 0.3092 | 1.0259 |
| Before pre fMRI | Negative score | 3.25 ± 1.95 | 2.84 ± 1.49 | 0.3717 | 0.9002 |
| After post fMRI | Control score | 1.98 ± 1.25 | 1.72 ± 1.15 | 0.4056 | 0.8377 |
| After post fMRI | Desire score | 1.92 ± 1.03 | 1.49 ± 0.81 | 0.0719 | 1.8331 |
| After post fMRI | Negative score | 2.73 ± 1.86 | 2.10 ± 1.56 | 0.1625 | 1.4148 |
| Day after | Control score | 1.70 ± 1.10 | 2.00 ± 1.38 | 0.3617 | -0.9197 |
| Day after | Desire score | 1.68 ± 0.82 | 1.34 ± 0.50 | 0.0613 | 1.9096 |
| Day after | Negative score | 2.17 ± 1.51 | 1.92 ± 1.06 | 0.4773 | 0.7155 |

### S10. Interaction with the insula during cue reactivity

As an exploratory analysis, we examined the interaction with insula subregions in addition to the striatum and amygdala. When subcortical targets were included in the model, two insular subregions—dorsal angula (β = 0.22) and ventral angula (β = 0.19)—showed uncorrected p-values < 0.05. Both regions exhibited increased PPI with all frontoparietal nodes, except the right frontal node, which showed decreased connectivity.

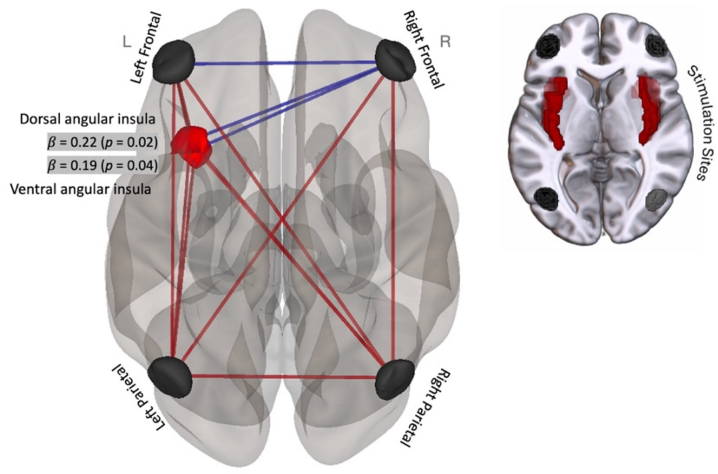


**Figure S5. Frontoparietal interactions with insular cortex.** Exploratory analysis of insula subregions revealed significant modulation of PPI connectivity with frontoparietal nodes during stimulation. Both the dorsal (β = 0.22, p = 0.02) and ventral (β = 0.19, p = 0.04) angular insula showed increased PPI with all frontoparietal regions except the right frontal node, which exhibited decreased connectivity. Stimulation sites are shown in black, with affected insula subregions highlighted in red. Red and blue lines indicate increased and decreased connectivity, respectively.

### S11. Resting state connectivity

Resting-state fMRI showed a similar pattern of increased frontoparietal connectivity and decreased connectivity to the left medial amygdala, though these effects did not reach statistical significance.

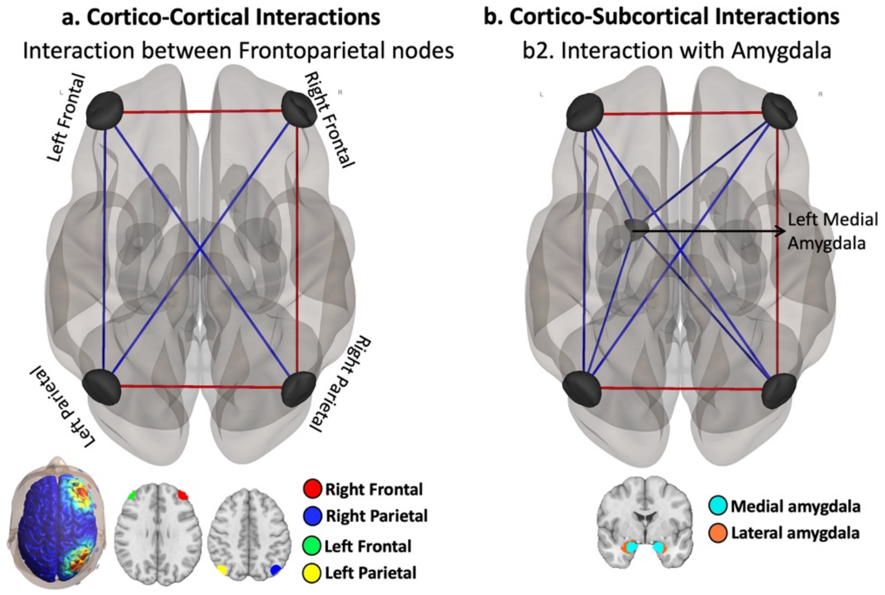


**Figure S6. Resting-State Connectivity Patterns Across Cortico-Cortical and Cortico-Subcortical Regions**. (a) Cortico-cortical interactions among frontoparietal nodes and (b) cortico-subcortical interactions between frontoparietal nodes and the left medial amygdala are shown. Red lines indicate increased functional connectivity, and blue lines indicate decreased connectivity following stimulation in the active group relative to sham.

### S12. Between-subject variations in frontoparietal connectivity

Figures S7 and S8 provide complementary visualizations of task-dependent gPPI connectivity effects during drug cue exposure for cortical and subcortical region pairs that exhibited significant time × group interactions in the main analyses. These figures illustrate individual-level pre- to post-assessment gPPI values and corresponding post–pre changes for both the active stimulation and sham groups, enabling inspection of the direction and distribution of stimulation-related effects. Individual participants exhibited variability in the magnitude and sign (positive vs. negative) of gPPI values, indicating interindividual differences in the direction and valence of connectivity changes following stimulation.

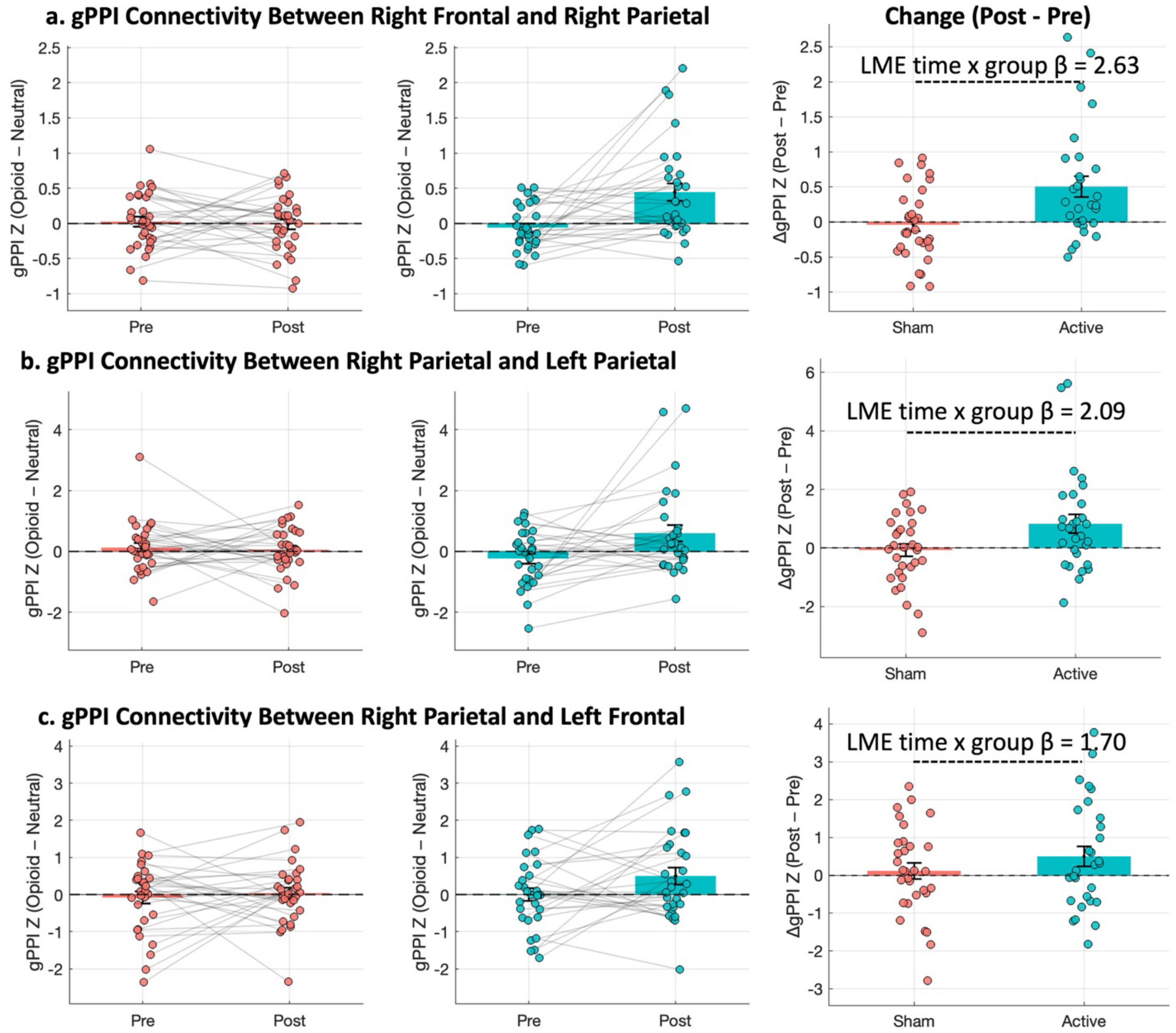


**Figure S7. Task-dependent gPPI connectivity changes during cue exposure for brain regions that showed significant positive time by group interactions.** (a) gPPI connectivity between the right frontal and right parietal regions, (b) right parietal and left parietal regions, and (c) right parietal and left frontal regions during drug cue exposure relative to neutral cues. Left and middle panels show individual pre- to post-assessment gPPI values for the sham and active stimulation groups, respectively, with lines connecting within-subject measurements. Right panels depict changes in gPPI values (Post − Pre) for each group. Points represent individual participants, bars indicate group means ± SEM, and the dashed horizontal line denotes zero (no task-dependent coupling). Positive values reflect stronger task-dependent functional coupling, whereas negative values reflect reduced or anticorrelated coupling. Reported β--values correspond to time × group interaction effects estimated using linear mixed-effects models.

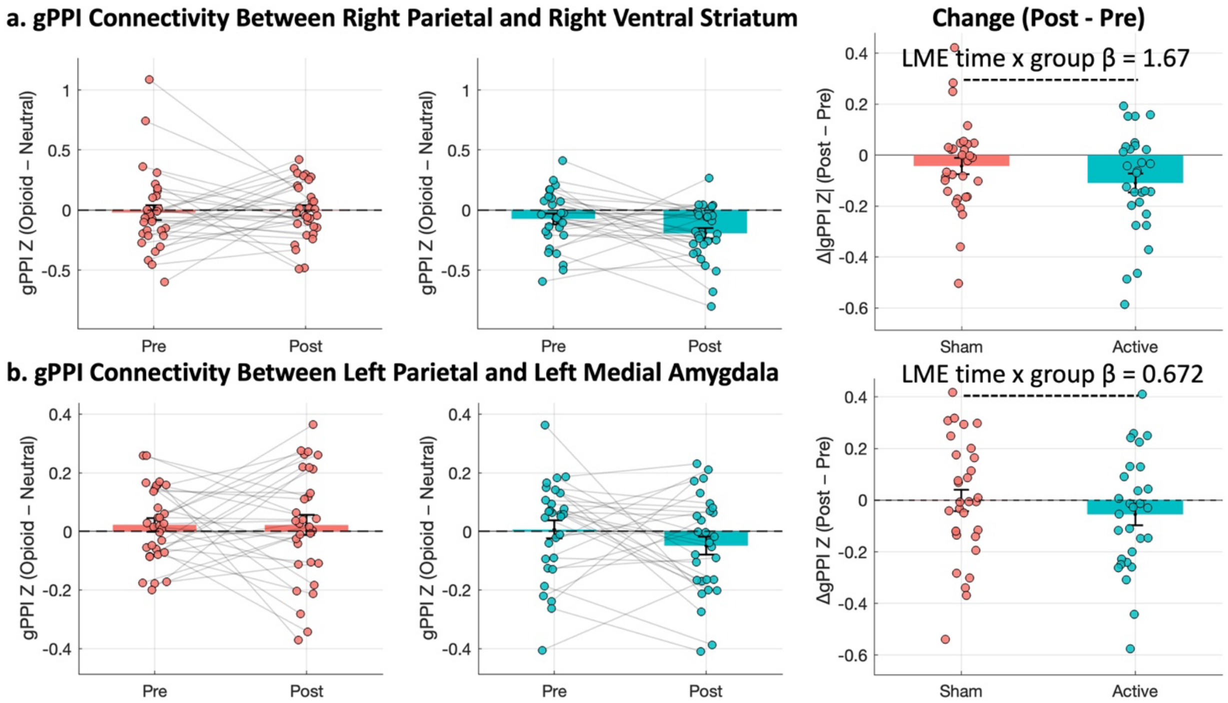


**Figure S8. Task-dependent gPPI connectivity between parietal and subcortical regions during cue exposure.** (a) gPPI connectivity between the right parietal cortex and right ventral striatum and (b) between the left parietal cortex and left medial amygdala during drug cue exposure relative to neutral cues. Left and middle panels show individual pre- to post-assessment gPPI values for the sham and active stimulation groups, respectively, with lines connecting within-subject measurements. Right panels depict changes in gPPI values (Post − Pre) for each group. Points represent individual participants, bars indicate group means ± SEM, and the dashed horizontal line denotes zero (no task-dependent coupling). Reported β-values correspond to time × group interaction effects estimated using linear mixed-effects models.

### S13. Inside scanner performance: reaction time and craving rating

#### S13.a. Response time to yellow border

Reaction time to the yellow border was analyzed using a linear mixed-effects model with Group (Active vs Sham), Time (Pre vs Post), and Condition (Cue vs Neutral) as fixed effects and a random intercept for subject. A significant main effect of Condition was observed ($F(1,182) = 7.52$, $p = 0.0067$), indicating slower responses during drug cue blocks compared with neutral blocks. Specifically, reaction times during neutral blocks were on average 0.31 s faster than during drug blocks ($\beta = -0.31$ s, 95% CI [−0.53, −0.09]). No main effects of Time ($F(1,182) < 0.001$, $p = 1.00$) or Group ($F(1,182) = 3.17$, $p = 0.076$) were detected. Furthermore, no significant interactions were observed between Group and Time ($p = 1.00$), Group and Condition ($p = 0.41$), Time and Condition ($p = 1.00$), or Group × Time × Condition ($p = 1.00$), indicating that stimulation did not modulate reaction time either overall or specifically during cue exposure.

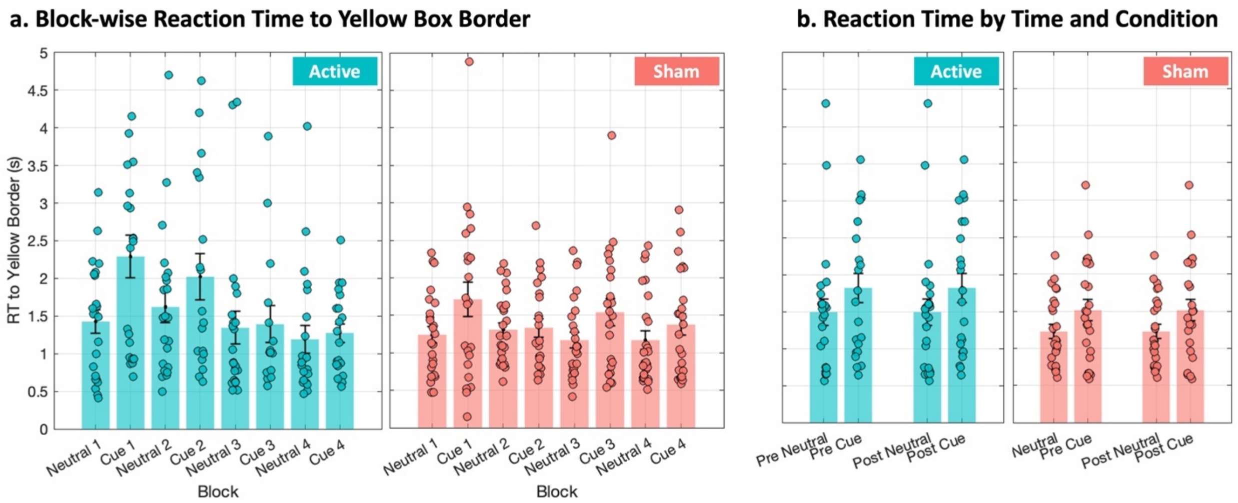


**Figure S9. Reaction time to the yellow box border during the cue reactivity task.** (a) Block-wise reaction time (RT; seconds) to the yellow box border during neutral and cue blocks, shown separately for the active (left) and sham (right) groups. Odd-numbered blocks correspond to neutral blocks and even-numbered blocks correspond to cue blocks. (b) Mean reaction time by Time (Pre vs Post) and Condition (Neutral vs Cue) for the active and sham groups. Bars represent group means ± SEM, and dots indicate individual participants. Reaction times were slower during cue blocks compared with neutral blocks, with no significant effects of Time or Group.

### S13.b. Craving rating inside the scanner

Craving ratings were analyzed using a linear mixed-effects model with Group (Active vs Sham), Time (Pre vs Post), and Condition (Cue vs Neutral) as fixed effects and a random intercept for subject. A significant main effect of Condition was observed ($F(1,188) = 10.38$, $p = 0.0015$), with higher craving ratings during cue blocks compared with neutral blocks ($\Delta \approx 0.45$ Likert units). No main effects of Time ($p = 1.00$) or Group ($p = 0.58$) were detected. A trend-level Group × Condition interaction was observed ($F(1,188) = 3.74$, $p = 0.054$), reflecting a numerically smaller cue–neutral craving difference in the active group relative to sham; however, this effect did not reach statistical significance. No interactions involving Time were significant (all $p = 1.00$), indicating that craving responses were stable across sessions (Figure S10).

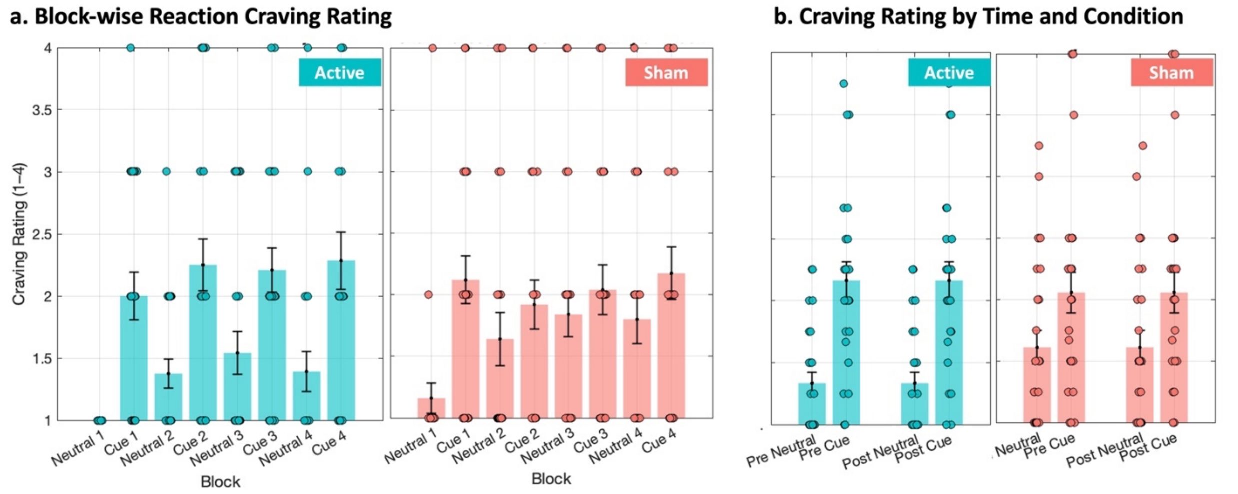


**Figure S10. Craving ratings during the cue reactivity task.** (a) Block-wise craving ratings (Likert scale, 1–4) during neutral and cue blocks, shown separately for the active (left) and sham (right) groups. Odd-numbered blocks correspond to neutral blocks and even-numbered blocks correspond to cue blocks. (b) Mean craving ratings by Time

(Pre vs Post) and Condition (Neutral vs Cue) for the active and sham groups. Bars represent group means ± SEM, and dots indicate individual participants.

#### S13.c. Modelling the yellow border component in functional connectivity analyses

To evaluate the potential influence of the yellow-border presentation on task-related activation estimates, we explicitly modeled the timing of the yellow-border events for each subject as a separate regressor in the first-level general linear model (GLM). As shown in Figure S10 for a sample subject in post-fMRI.

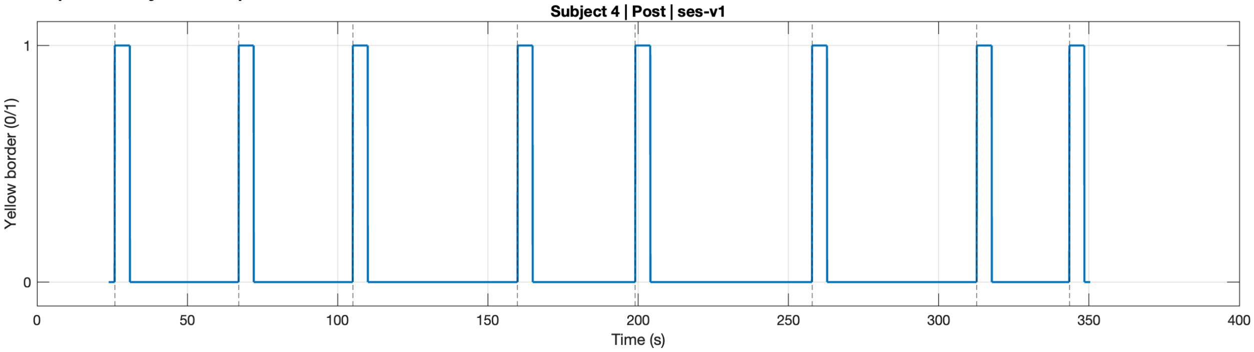


**Figure S10. Example time series of the yellow-border (BOX_ONSET) regressor for a single participant (Subject 4, Post).** The binary trace indicates periods when the yellow border was present (value = 1) and absent (value = 0).

Yellow-border onsets were entered as an event-related regressor and convolved with the canonical hemodynamic response function, thereby accounting for the delayed and temporally extended hemodynamic response associated with these events. This regressor was included as a regressor of no interest, distinct from the drug cue and neutral cue regressors, to ensure that variance associated with attentional engagement or response monitoring related to the border presentation was not attributed to cue-related neural activity. All other aspects of the GLM—including cue and neutral condition regressors, motion parameters, and standard nuisance regressors—were held constant across models. Because the yellow-border events were explicitly modeled with HRF convolution, no additional exclusion or censoring of time points following the border presentation was applied, as the hemodynamic carryover associated with these events was appropriately captured within the model. To assess robustness, task-related activation results were compared between models with and without inclusion of the yellow-border regressor for both pre- and post-stimulation fMRI sessions.

#### S13.d. Modelling the yellow border component in functional activity analyses

Opioid and neutral cue blocks were modeled using sustained block regressors (BLOCK(31,1)), and the yellow-box component was modeled using a shorter block regressor (BLOCK(5,1)). The primary contrast (Opioid > Neutral) was estimated while controlling for variance associated with the yellow-box regressor. All regressors were convolved within a single GLM framework, which accounts for the temporal overlap of the hemodynamic response across adjacent task components. Baseline and low-frequency trends were estimated implicitly, and the first three TRs were removed to minimize non–steady-state effects. Including the yellow-box regressor did not alter the pattern of task-related activation or connectivity results. You can see the results for one sample subject in the pre-stimulation session. We did this analysis for the pre-stimulation session, and group-level results are similar to what we presented in the main manuscript.

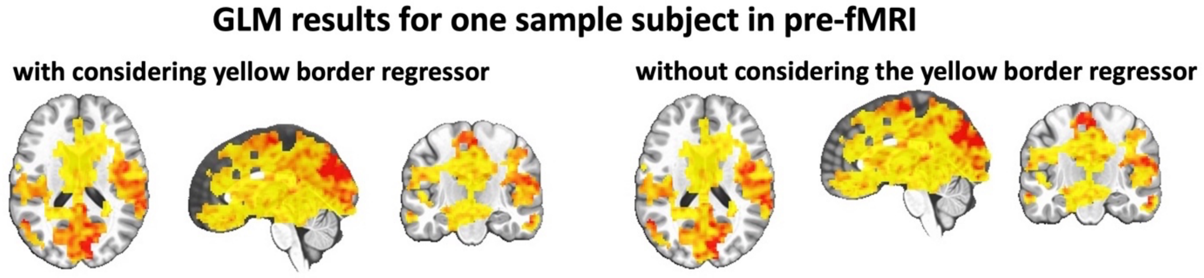


**Figure S11.** Representative single-subject GLM results from the pre-fMRI session are shown with (left) and without (right) inclusion of the yellow-border presentation as a regressor of no interest.

### S14. Abstinence Duration

Abstinence duration and recent substance use were assessed at baseline using the Participant Last Use Summary (PLUS) questionnaire. We summarized days since last illicit substance use, along with alcohol, tobacco, and caffeine use measures, and visualized their distributions for the sham and active groups using bar plots with individual data points. Abstinence duration exhibited substantial inter-individual variability beyond the minimum abstinence requirement. We additionally examined whether baseline abstinence duration and related substance-use variables were associated with cue-induced craving or moderated responses to the applied stimulation; no predictive/correlative relationships were observed.

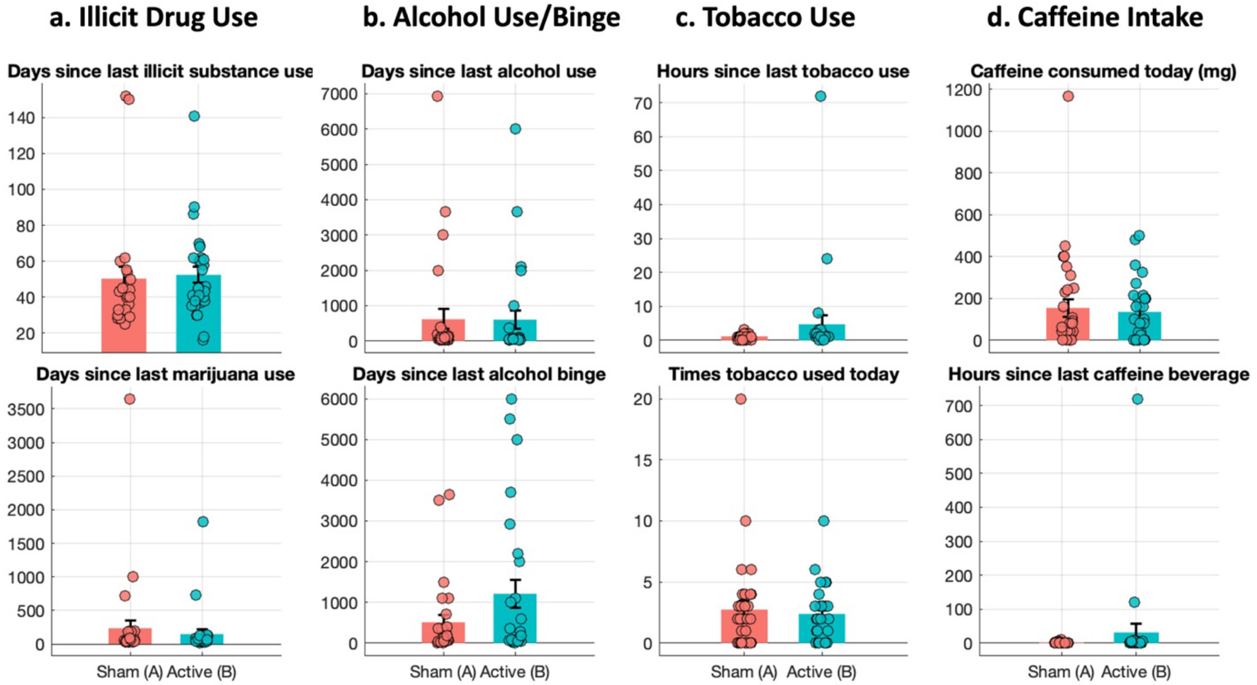


**Figure S12. Baseline substance use and abstinence characteristics assessed with the Participant Last Use Summary (PLUS).** Baseline distributions of substance use–related variables are shown for the sham (in red) and active (in blue) groups at the pre-stimulation assessment. Panels depict days since last illicit substance use and marijuana use (a), days since last alcohol use and binge episodes (b), tobacco use (hours since last use and number of uses on the assessment day; c), and caffeine intake (consumed dose and hours since last caffeinated beverage; d). Bars represent group means with standard error of the mean, and overlaid points show individual participant values.

**Table S5. Baseline abstinence duration and substance use characteristics in the Sham group**

| | N | Median | Mean | SD | p_value | z_value |
|---|---|---|---|---|---|---|
| Days since last marijuana use | 30 | 50 | 234.516 | 666.802 | 0.779 | -0.281 |
| Times tobacco used today | 30 | 2 | 2.742 | 3.941 | 0.804 | -0.249 |
| Hours since last tobacco use | 28 | 1 | 1.071 | 0.858 | 0.507 | -0.664 |
| Days since last alcohol use | 25 | 60 | 353.800 | 903.411 | 0.846 | -0.194 |
| Days since last alcohol binge | 25 | 124 | 566.880 | 985.872 | 0.479 | -0.709 |
| Days since last illicit substance use | 24 | 42 | 50.250 | 32.611 | 0.214 | -1.243 |
| Days since last medication use | 22 | 1 | 0.773 | 0.528 | 0.937 | -0.079 |
| Caffeine consumed today (mg) | 30 | 60 | 154.161 | 232.285 | 0.870 | 0.164 |
| Hours since last caffeine beverage | 30 | 1 | 1.333 | 1.988 | 0.051 | -2.045 |

**Table S6. Baseline abstinence duration and substance use characteristics in the Active group**

| | N | Median | Mean | SD | p_value | z_value |
|---|---|---|---|---|---|---|
| Days since last marijuana use | 28 | 60 | 148.214 | 353.295 | 0.779 | -0.281 |
| Times tobacco used today | 29 | 2 | 2.379 | 2.274 | 0.804 | -0.249 |
| Hours since last tobacco use | 28 | 1 | 4.679 | 13.952 | 0.507 | -0.664 |
| Days since last alcohol use | 25 | 61 | 411.400 | 882.777 | 0.846 | -0.194 |
| Days since last alcohol binge | 25 | 186 | 1064.680 | 1606.952 | 0.479 | -0.709 |
| Days since last illicit substance use | 29 | 45 | 52.414 | 24.680 | 0.214 | -1.243 |
| Days since last medication use | 24 | 1 | 1.042 | 1.429 | 0.937 | -0.079 |
| Caffeine consumed today (mg) | 29 | 100 | 134.793 | 144.701 | 0.870 | 0.164 |
| Hours since last caffeine beverage | 29 | 2 | 30.966 | 134.328 | 0.051 | -2.045 |

### S15. Inter-individual variability and pre/post values in functional activity

To characterize **inter-individual variability and group-level functional activity** in subcortical regions, functional activation was extracted for each participant at **pre- and post-stimulation** time points and summarized separately for the **Active** and **Sham** groups. Atlas-based parcellation was performed using **Brainnetome atlas subcortical regions**, including the **medial amygdala (mAmyg), lateral amygdala (lAmyg), rostral hippocampus (rHipp), caudal hippocampus (cHipp), ventral caudate (vCa), globus pallidus (GP), nucleus accumbens (NAC), ventromedial putamen (vmPu), dorsal caudate (dCa), and dorsolateral putamen (dlPu).** For each subregion and time point, group-level mean β values and standard errors are shown, with individual subject values overlaid to illustrate variability across participants. Data are presented separately for each group and time point, enabling visualization of subject-level heterogeneity and changes in functional activity across stimulation sessions (Figure S12).

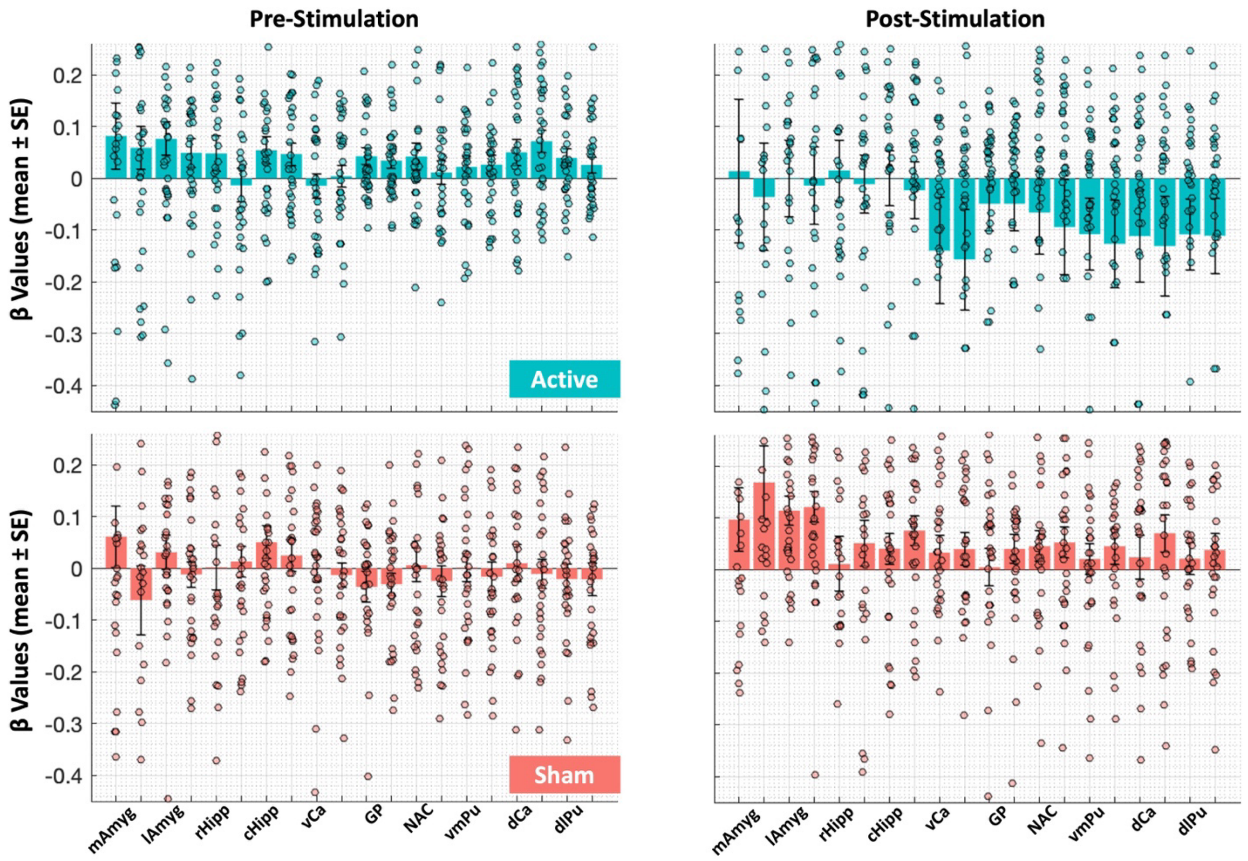


**Figure S13. Subcortical β values before and after stimulation in Active and Sham groups.** Bar plots show mean ± SE of β values across subjects for each subcortical region during **pre-stimulation** (left column) and **post-stimulation** (right column), separately for the **Active** (top row, teal) and **Sham** (bottom row, red) groups. Individual subject data points are overlaid as jittered dots. Regions include medial amygdala (mAmyg), lateral amygdala (lAmyg), rostral hippocampus (rHipp), caudal hippocampus (cHipp), ventral caudate (vCa), globus pallidus (GP), nucleus accumbens (NAC), ventromedial putamen (vmPu), dorsal caudate (dCa), and dorsolateral putamen (dlPu). For each subregion, the first bar corresponds to the left hemisphere and the second bar corresponds to the right hemisphere. Horizontal dashed lines indicate zero, and error bars represent the standard error of the mean.

Atlas-based parcellation was performed again using **Brainnetome atlas–defined cingulate subregions**, including **dorsal area 23 (A23d), ventral area 23 (A23v), and caudal area 23 (A23c)**—corresponding to subdivisions of the **posterior cingulate cortex**—as well as **rostroventral area 24 (A24rv), caudodorsal area 24 (A24cd), pregenual area 32 (A32p), and subgenual area 32 (A32sg)**, representing anterior and mid-cingulate regions (Figure S13).

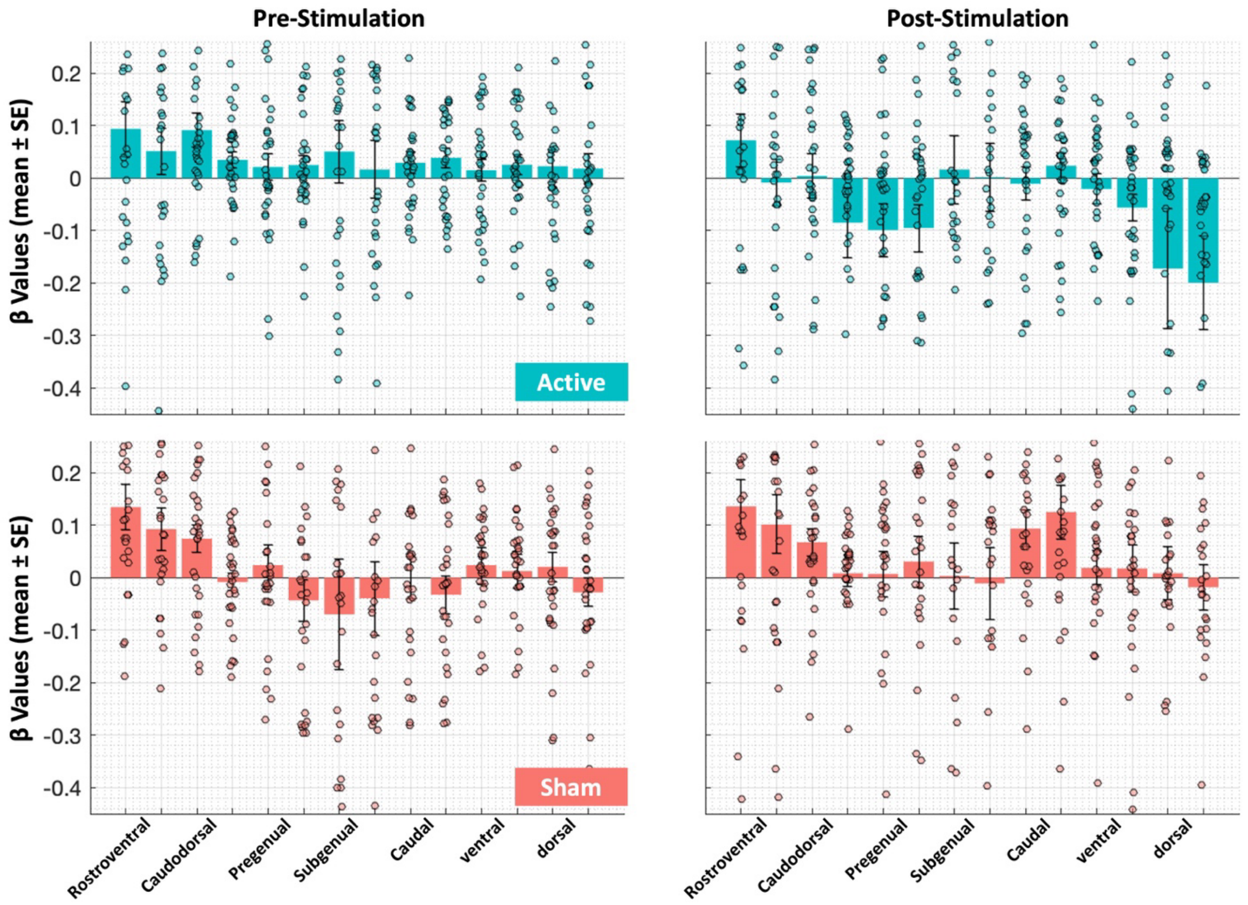


**Figure S14. Cingulate cortex β values before and after stimulation in Active and Sham groups.** Bar plots show mean ± SE of β values across subjects for each **Brainnetome atlas–defined cingulate subregion** during **pre-stimulation** (left column) and **post-stimulation** (right column), shown separately for the **Active** group (top row, teal) and the **Sham** group (bottom row, red). Individual subject data points are overlaid as jittered dots to illustrate inter-individual variability. Regions included in this analysis comprise **dorsal area 23 (A23d), ventral area 23 (A23v), caudal area 23 (A23c)**—corresponding to subdivisions of the **posterior cingulate cortex**—as well as **rostroventral area 24 (A24rv), caudodorsal area 24 (A24cd), pregenual area 32 (A32p), and subgenual area 32 (A32sg)**, corresponding to anterior and mid-cingulate subdivisions. For each subregion, the **first bar corresponds to the left hemisphere and the second bar corresponds to the right hemisphere.** Horizontal dashed lines indicate zero β values, and error bars represent the **standard error of the mean.**

Because striatal seed definition in the main analyses was based on a different atlas, we additionally replicated the analysis pipeline using anatomically defined striatal regions. Specifically, we applied the same pre- to post-stimulation analytical framework to striatal subregions defined a priori based on anatomical criteria, allowing us to visualize and compare within-subject changes in functional activity across time. This complementary analysis ensured that observed pre-to-post patterns were not dependent on atlas selection and provided a convergent anatomical perspective on striatal involvement.

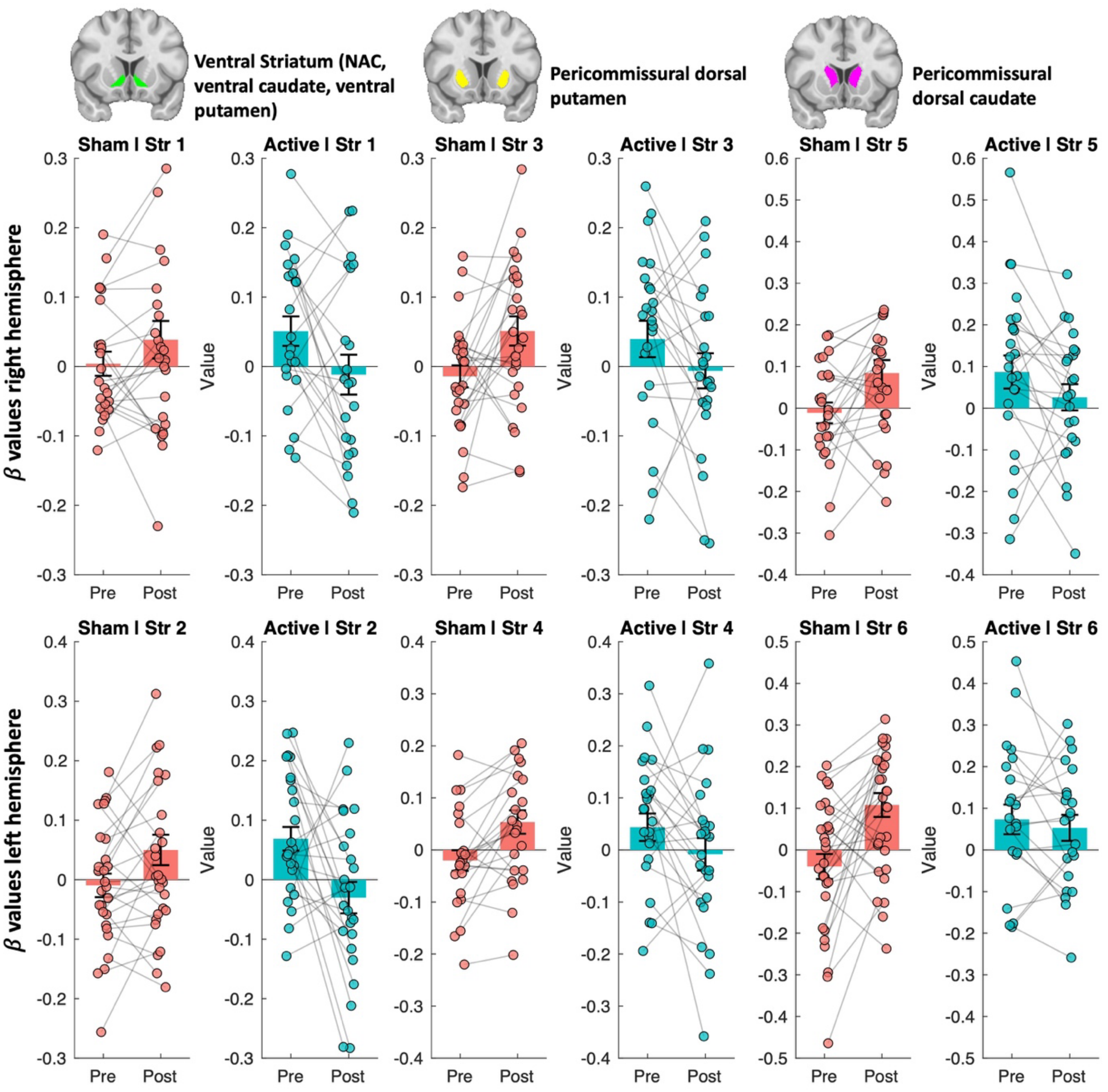


**Figure S15. Pre- to post-stimulation changes in striatal β values across sham and active groups.** Paired scatter and bar plots show individual subject β values extracted from six striatal subregions before (Pre) and after (Post) stimulation, displayed separately for the sham (red) and active (cyan) groups. Each dot represents a participant, with gray lines indicating within-subject changes across time. Bars represent group means and error bars denote ± SEM. Top row shows right-hemisphere striatal subregions; bottom row shows left-hemisphere subregions. Anatomical locations of the striatal regions are illustrated above each column, including ventral striatum (nucleus accumbens, ventral caudate, ventral putamen), pericommissural dorsal putamen, and pericommissural dorsal caudate.

### S16. Exploratory analysis on craving

To formally test whether these associations differed between groups, we fitted a linear model including a Group × connectivity interaction. This interaction did not reach statistical significance ($\beta = -68.73$, $p = 0.12$), indicating that the difference in slopes between the active and sham groups was not statistically reliable. Accordingly, these findings should be interpreted as exploratory. We also applied correlation analyses between VAS score changes and frontoparietal PPI connectivity at different time points, and we couldn't find significant (p uncorrected < 0.05) results (Figure S15).

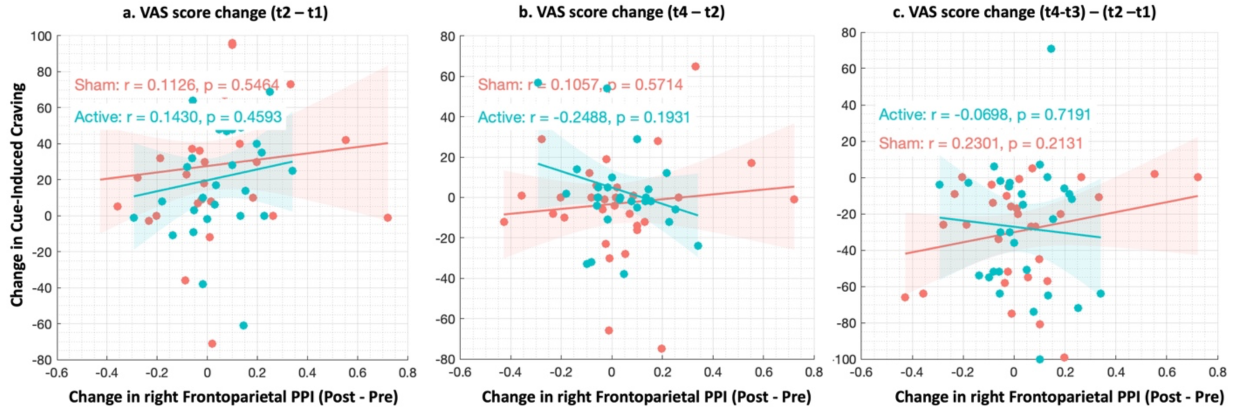


**Figure S16. Associations between changes in frontoparietal connectivity and craving across task and cue conditions.** Scatter plots show the relationship between changes in right frontoparietal psychophysiological interaction (PPI; post minus pre) and changes in craving measured using visual analog scales (VAS), plotted separately for the sham (red) and active (teal) stimulation groups. Panel (a) shows task-induced craving change before fMRI (t2 - t1). Panel (b) shows post-fMRI cue-induced craving change (t4 - t2). Panel (c) shows the difference-of-differences for task-induced craving, defined as the post-fMRI task change (t4 - t3) minus the pre-fMRI task change (t2 - t1). Solid lines represent linear regression fits for each group, with shaded areas indicating 95% confidence intervals. Pearson correlation coefficients and corresponding p-values are shown for each group in each panel.

#### S16.2. Cue-induced craving based on the VAS score at different time points

##### Cue-induced craving changes between post-fMRI assessments (t4 – t2)

Cue-induced craving decreased from the first to the second post-fMRI assessment in both groups. The sham group showed a mean change of −29.4 (SD = 28.2, SEM = 5.06), while the active stimulation group showed a mean change of −27.8 (SD = 34.9, SEM = 6.47). There was no significant difference between groups (Welch's t(53.9) = −0.20, p = 0.84), and the 95% confidence interval for the group difference ranged from −18.1 to 14.8, indicating substantial overlap between groups.

##### Task-induced craving changes before and after fMRI (t2 – t1)

Before tACS fMRI, task-induced craving increased in both groups, with a mean change of 28.1 (SD = 36.8, SEM = 6.60) in the sham group and 20.8 (SD = 30.2, SEM = 5.61) in the active group. After fMRI, task-induced craving was attenuated in both groups, with a mean change of −2.9 (SD = 26.6, SEM = 4.78) in the sham group and 3.4 (SD = 23.0, SEM = 4.28) in the active group. When task-induced craving changes were summarized using a difference-of-differences metric (post-fMRI task change minus pre-fMRI task change), both groups showed an overall reduction, with a larger mean decrease in the sham group (−31.0, SD = 45.9, SEM = 8.24) than in the active group (−17.3, SD = 37.8, SEM = 7.02). However, this between-group difference was not statistically significant (Welch's t(57.1) = −1.27, p = 0.21; 95% CI −35.4 to 8.0).

##### Linear mixed-effects analysis of task-induced craving

A linear mixed-effects model including Group, Task Interval (pre- vs post-fMRI), and their interaction, with a random intercept for participant, revealed a significant main effect of Task Interval (F(1,116) = 17.49, $p < 0.001$). Across both groups, task-induced craving was significantly lower after fMRI compared with before fMRI (estimate = −31.0, 95% CI −45.7 to −16.3). There was no significant main effect of Group (F(1,116) = 0.94, p = 0.33), and the Group × Task Interval interaction was not significant (F(1,116) = 1.65, p = 0.20), indicating that the magnitude of the pre- to post-fMRI change in task-induced craving did not differ significantly between the sham and active stimulation groups.

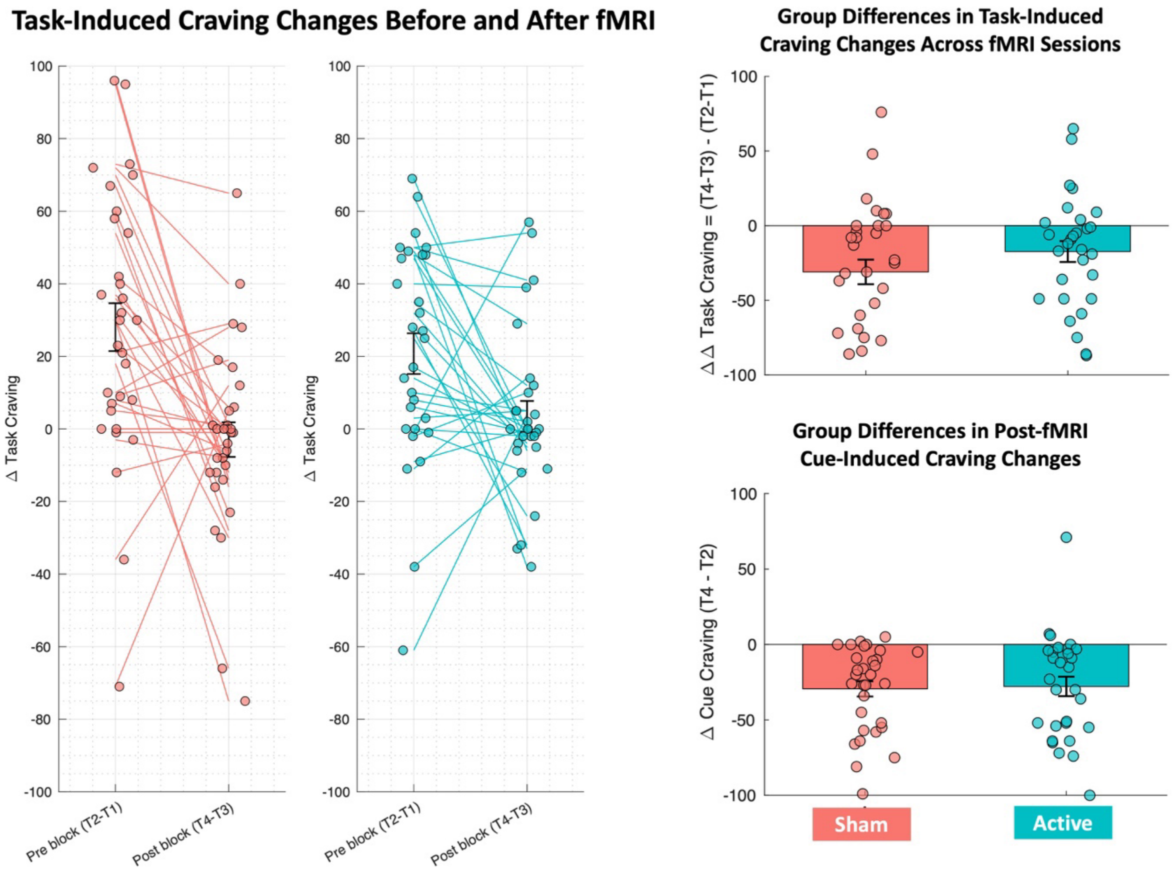


**Figure S17. Task- and cue-induced craving changes before and after fMRI.** Left panels show individual within-subject trajectories of task-induced craving changes measured before fMRI (t2 - t1) and after fMRI (t4 - t3), plotted separately for the sham and active stimulation groups. Lines connect paired pre- and post-fMRI measurements within each participant. The top-right panel summarizes group differences in task-induced craving using a difference-of-differences approach, defined as the post-fMRI task-induced change (t4 - t3) minus the pre-fMRI task-induced change (t2 - t1), which accounts for within-subject variability across sessions. The bottom-right panel shows group differences in post-fMRI cue-induced craving change, defined as t4 - t2. Bars represent mean ± SEM, with individual participant values overlaid.

#### S16.3. Correlation analyses between VAS score changes and functional activity

To examine the relationship between regional functional brain activity and subjective craving, exploratory correlation analyses were conducted within the active stimulation group. Functional activation values were extracted for each participant from 246 Brainnetome Atlas subregions, yielding subject-by-ROI matrices for pre and post fMRI. For each craving metric, ROI-wise correlations were computed between craving values and regional BOLD activation changes across participants using Pearson correlation coefficients. Analyses were restricted to the active stimulation group to assess brain–behavior relationships following active neuromodulation. To reduce the influence of extreme values, bivariate outliers were identified using a robust median absolute deviation (MAD) criterion applied jointly to craving and BOLD values. Observations flagged as outliers in either dimension were excluded before correlation estimation. Correlation coefficients and associated p-values were then recomputed on the cleaned data. For visualization, excluded outliers were plotted separately to ensure transparency. Given the exploratory nature of these secondary analyses and the large number of regional tests, statistical significance was first evaluated at an uncorrected threshold of $p < 0.05$ to identify nominal associations. In parallel, false discovery rate (FDR) correction using the Benjamini–Hochberg procedure was applied across the 246 Brainnetome subregions for each craving metric to account for multiple comparisons. Scatter plots were generated for selected regions of interest to illustrate the relationship between craving measures and functional activity changes, with regression lines fitted to the non-outlier data.

As shown in Figure S16, across these subregions, predominantly negative associations indicate that lower post-stimulation craving is associated with greater BOLD down-regulation, with the most robust effects observed in striatal components of the basal ganglia, particularly the nucleus accumbens and caudate subregions**.**

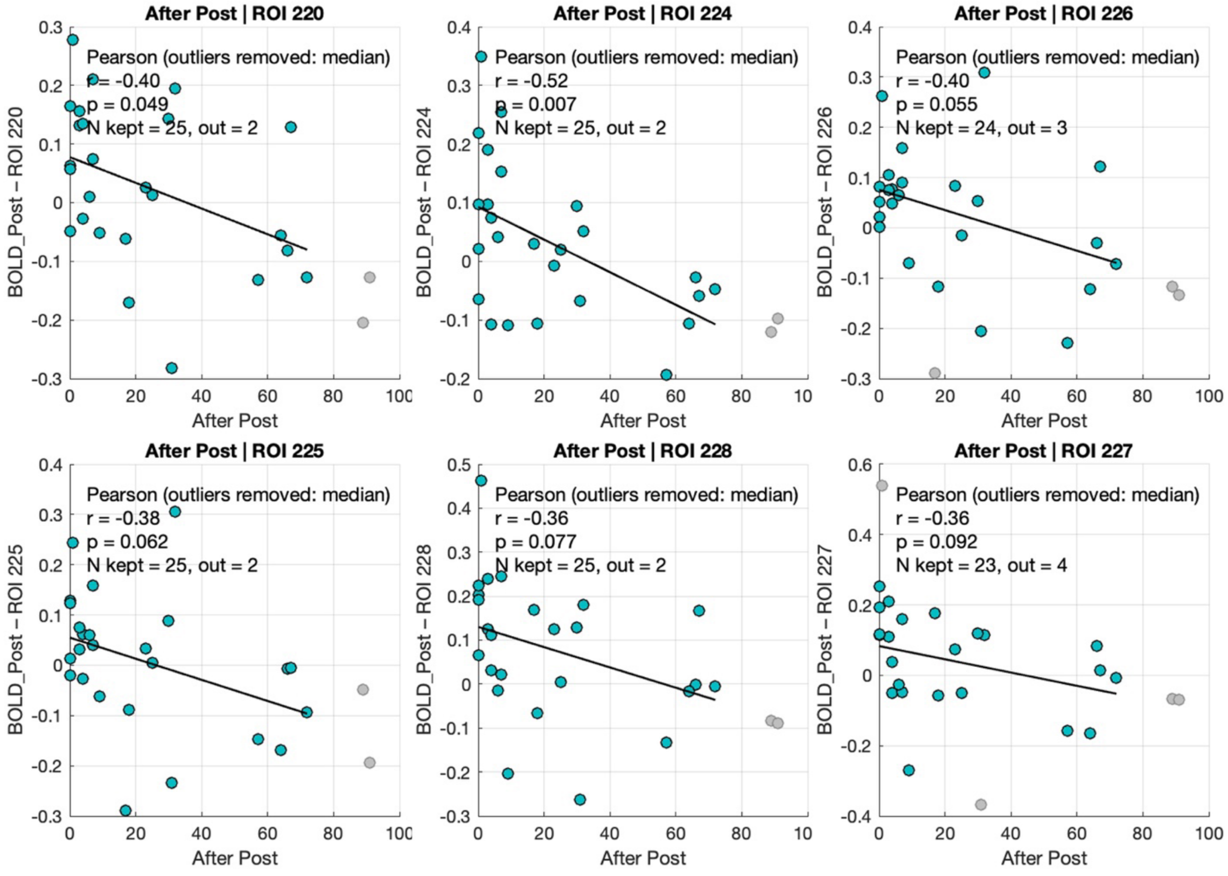

**Figure S18. BOLD-Craving correlations across subcortical subregions after stimulation.** Scatter plots illustrate exploratory Pearson correlations between post-stimulation craving (After fMRI Post Stimulation) and post-stimulation BOLD signal change across subcortical Brainnetome Atlas (BNA) ROIs spanning indices 211–230. Each point represents an individual participant; solid lines denote least-squares regression fits. Correlation coefficients (r), p-values, and sample sizes (N) are reported within each panel. Outliers were identified using a median-based criterion and are shown in gray when excluded. The analyzed subregions include the amygdala (medial amygdala: ROIs 211–212; lateral amygdala: ROIs 213–214), hippocampus (rostral hippocampus: ROIs 215–216; caudal hippocampus: ROIs 217–218), and basal ganglia (ventral caudate: ROIs 219–220; globus pallidus: ROIs 221–222; nucleus accumbens: ROIs 223–224; ventromedial putamen: ROIs 225–226; dorsal caudate: ROIs 227–228; dorsolateral putamen: ROIs 229–230).

### S16.4. Correlation analyses between VAS score changes and electric field

A similar approach was used to examine correlations between tACS-induced electric field strength and changes in craving scores across different time points. Brainnetome atlas parcellation was applied to individualized head models, and electric field strength was extracted from 210 cortical regions. Correlation analyses were restricted to regions receiving an electric field strength of at least 0.06 V/m to exclude off-target brain areas.

When an outlier-removal approach was applied, following the procedure described in Section S16.3, no correlations survived an uncorrected threshold of $p < 0.05$. However, prior to outlier removal, exploratory correlation analyses within the active stimulation group revealed that greater electric field strength in inferior parietal lobule subregions was associated with greater reductions in craving. Specifically, cue-induced craving change following stimulation (T4 − T2) was negatively correlated with electric field magnitude in angular and supramarginal gyrus subregions (A39c/PGp, A39rd/Hip3, A39rv/PGa, A40rd/PFt; $r = -0.43$ to $-0.53$, all $p < 0.05$, uncorrected). In addition, electric field strength in A39rd and A40c subregions was associated with reduced task-induced craving after stimulation and with task-related difference-in-differences metrics, indicating stimulation-specific modulation of craving during cognitive engagement.

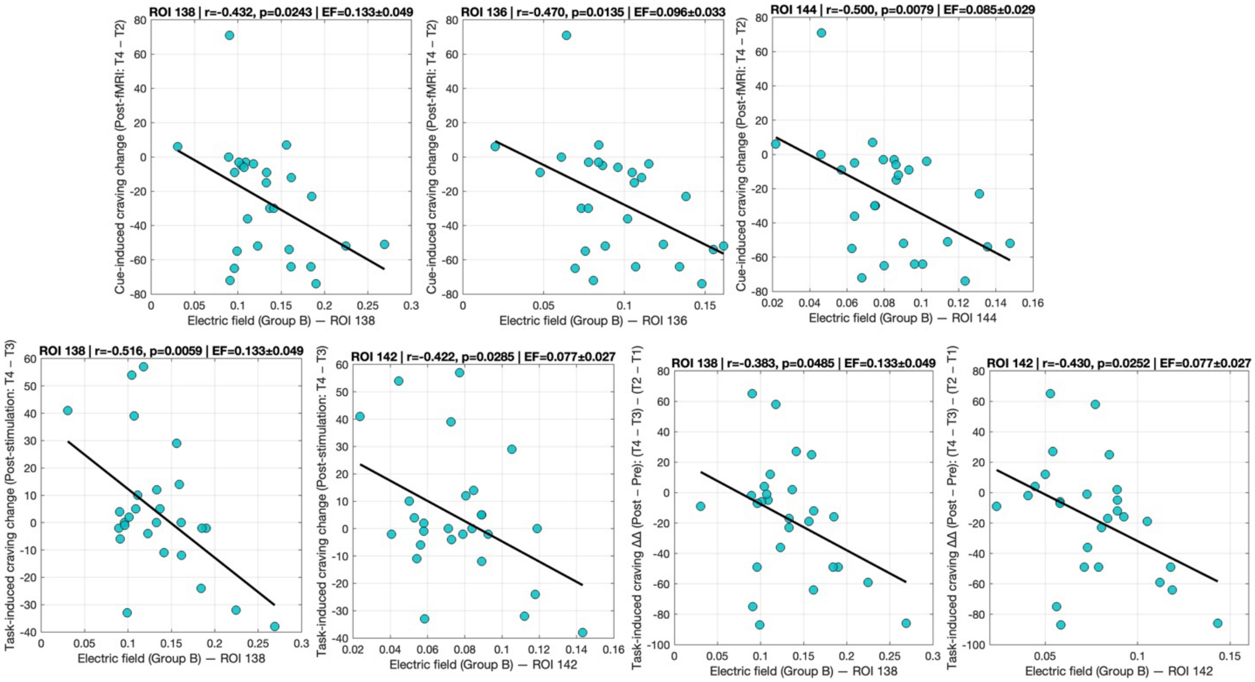

**Figure S19. Exploratory associations between tACS-induced electric field strength and craving change in the active stimulation group.** Scatterplots depict uncorrected correlations between regional electric field (EF) magnitude and changes in craving across different time points. **Top row** shows cue-induced craving change following stimulation (T4 − T2) as a function of EF strength in inferior parietal lobule subregions, including ROI 138 (A39rd, rostrodorsal area 39; Hip3)**,** ROI 136 (A39c, caudal area 39; PGp)**,** and ROI 144 (A39rv, rostroventral area 39; PGa)**.** **Bottom row** illustrates associations between EF strength and task-induced craving change after stimulation (T4 − T3) and task-related difference-in-differences [(T4 − T3) − (T2 − T1)] in ROI 138 (A39rd, Hip3) and ROI 142 (A40c, caudal area 40; PFm). Each point represents an individual participant, and solid lines indicate least-squares regression fits. Across panels, higher EF magnitude in angular and supramarginal gyrus subregions was associated with greater reductions in craving. All results are shown without outlier removal to preserve inter-individual variability in delivered stimulation dose and are reported at an uncorrected threshold ($p < 0.05$).

### S17. Brief Craving Scale

Across all four BCS items (intensity, frequency, duration, and number of craving episodes), no significant differences were observed between the sham and active stimulation groups at baseline**,** before pre-fMRI, or day-after assessments (Welch's t-tests, all $p > 0.05$; Table S7).

**Table S7. Brief Craving Scale at three different time points**

| BCS_Label | Timepoint | Mean_Sham | SEM_Sham | Mean_Active | SEM_Active | t | df | p_ttestWelch | p_ranksum | Hedges'g |
|---|---|---|---|---|---|---|---|---|---|---|
| BCS-1: Intensity (0-4) | Baseline | 1.032 | 0.170 | 1.483 | 0.162 | -1.918 | 57.983 | 0.060 | 0.034 | 0.496 |
| BCS-1: Intensity (0-4) | Before pre-fMRI | 1.032 | 0.170 | 1.310 | 0.150 | -1.226 | 57.498 | 0.225 | 0.151 | 0.316 |
| BCS-1: Intensity (0-4) | Day after | 0.800 | 0.162 | 0.857 | 0.143 | -0.265 | 55.563 | 0.792 | 0.598 | 0.069 |
| BCS-2: Frequency (0-4) | Baseline | 1.129 | 0.172 | 1.517 | 0.137 | -1.766 | 56.005 | 0.083 | 0.068 | 0.454 |
| BCS-2: Frequency (0-4) | Before pre-fMRI | 1.226 | 0.159 | 1.517 | 0.128 | -1.431 | 56.189 | 0.158 | 0.123 | 0.368 |
| BCS-2: Frequency (0-4) | Day after | 0.833 | 0.152 | 0.893 | 0.149 | -0.280 | 55.993 | 0.781 | 0.702 | 0.074 |
| BCS-3: Duration (0-4) | Baseline | 1.161 | 0.197 | 1.414 | 0.136 | -1.054 | 52.584 | 0.297 | 0.161 | 0.270 |
| BCS-3: Duration (0-4) | Before pre-fMRI | 1.129 | 0.166 | 1.310 | 0.150 | -0.812 | 57.752 | 0.420 | 0.271 | 0.210 |
| BCS-3: Duration (0-4) | Day after | 0.800 | 0.139 | 0.821 | 0.127 | -0.114 | 55.815 | 0.910 | 0.846 | 0.030 |
| BCS-4: Count | Baseline | 7.548 | 3.320 | 3.552 | 0.578 | 1.186 | 31.814 | 0.244 | 0.550 | -0.298 |
| BCS-4: Count | Before pre-fMRI | 8.000 | 4.767 | 3.586 | 0.554 | 0.920 | 30.809 | 0.365 | 0.698 | -0.231 |
| BCS-4: Count | Day after | 3.167 | 0.844 | 2.714 | 0.678 | 0.418 | 54.242 | 0.678 | 0.544 | -0.109 |

### S18. Addressing baseline self-regulatory differences in a between-subjects design

Given the between-subjects design, we additionally examined cue-elicited executive control and disinhibition using the Cue-Elicited Experience (CEE) Drugs questionnaire. The CEE includes subscales indexing attentional capture, successful and unsuccessful effortful control, cognitive reappraisal, rumination, and metacognitive awareness during drug cue exposure. No significant between-group differences were observed on these executive control–related CEE subscales at baseline or post-stimulation assessments (all p's > 0.05; Table S8), indicating comparable cue-elicited self-regulatory capacity between the sham and active groups. Together with the absence of baseline differences in craving severity measured by the Brief Drug Craving Scale (Welch's t-tests, all $p > 0.05$; Table S7-baseline), these findings reduce the likelihood that observed group effects reflect pre-existing differences unrelated to the stimulation manipulation.

**Table S8. CEE Drug Questionnaire at baseline**

| Measure | Mean_Sham | SD_Sham | Mean_Active | SD_Active | p_ttestWelch | t | p_ranksum | Hedges' g |
|---|---|---|---|---|---|---|---|---|
| success_control_score | 212.968 | 114.245 | 247.172 | 101.354 | 0.224 | -1.229 | 0.147 | 0.317 |
| unsuccess_control_score | 164.645 | 103.747 | 215.103 | 114.816 | 0.080 | -1.782 | 0.077 | 0.464 |
| reappraisal_score | 209.677 | 114.207 | 264.793 | 110.924 | 0.063 | -1.896 | 0.030 | 0.491 |
| attentional_score | 258.516 | 113.142 | 274.448 | 107.268 | 0.578 | -0.560 | 0.620 | 0.145 |
| ruminative_score | 282.129 | 114.287 | 280.862 | 109.276 | 0.965 | 0.044 | 0.953 | -0.011 |
| gen_memory_score | 94.484 | 34.425 | 103.621 | 40.341 | 0.351 | -0.941 | 0.110 | 0.245 |
| interoceptive_score | 262.452 | 102.429 | 294.000 | 82.692 | 0.193 | -1.317 | 0.196 | 0.339 |
| metacog_score | 294.484 | 95.212 | 347.552 | 67.915 | 0.016 | -2.498 | 0.040 | 0.641 |
| neg_emotional_score | 206.871 | 117.244 | 270.586 | 103.510 | 0.029 | -2.235 | 0.020 | 0.577 |
| neg_memory_score | 146.581 | 91.166 | 196.276 | 74.627 | 0.024 | -2.317 | 0.038 | 0.597 |
| pos_appetitive_score | 165.677 | 128.532 | 147.931 | 120.638 | 0.583 | 0.552 | 0.610 | -0.143 |
| pos_memory_score | 82.677 | 76.116 | 62.172 | 57.636 | 0.243 | 1.181 | 0.459 | -0.303 |
| verbalizing_score | 271.677 | 89.395 | 273.276 | 72.309 | 0.939 | -0.076 | 0.525 | 0.020 |

### S19. Medication History

Medication use was assessed using a structured Medication Use History questionnaire administered at t0 (before pre-fMRI). Participants were asked to report all current medications taken within the past three months, including both prescription and non-prescription medications. Participants first indicated the total number of medications they were currently taking (range: 0–15). For each reported medication, detailed information was collected, including the medication name, dosage amount, dosage unit, frequency of use, and the time the medication was last taken. Space was also provided for additional notes to capture relevant medications or clarifying information not otherwise specified.

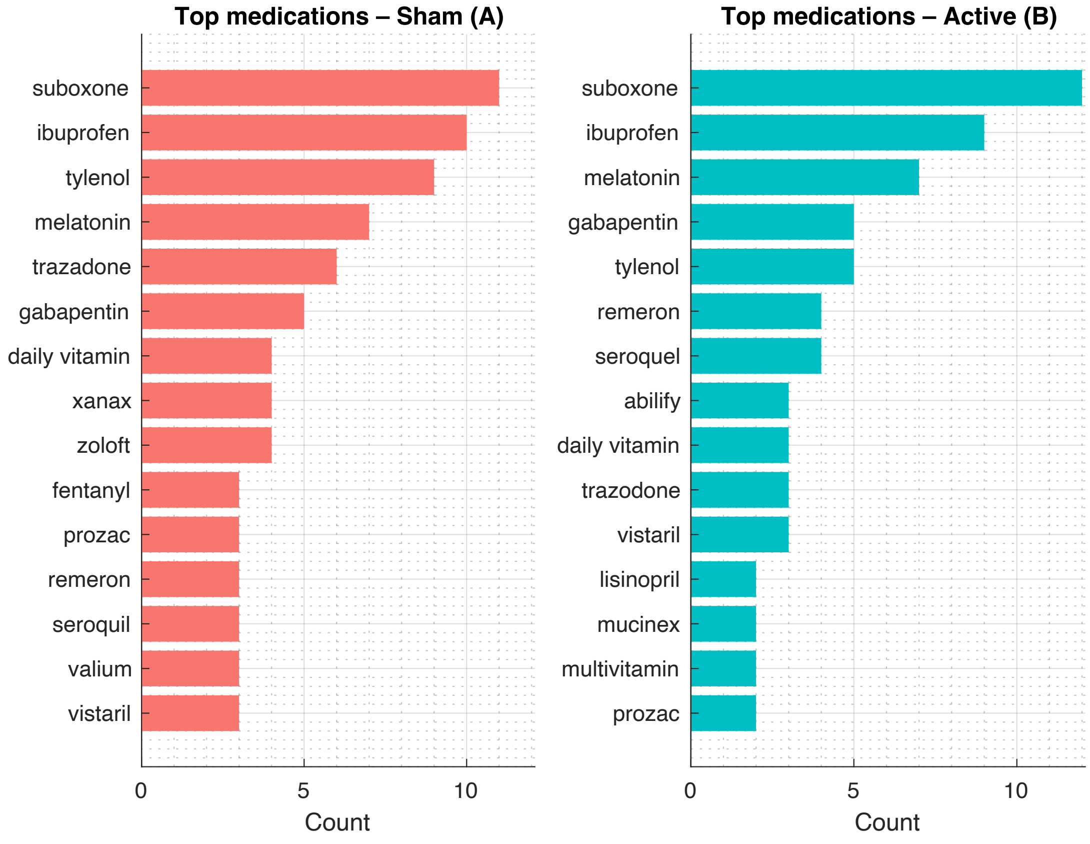


**Figure S20.** Most frequently reported medications in the sham and active groups at baseline. Bars indicate the number of participants reporting each medication at the before-pre-fMRI assessment.

**S20. Details about the p-value correction methods in each set of analysis**

**Functional activity**

Voxelwise statistical maps were thresholded at an uncorrected voxel-level threshold of $p < 0.005$, and multiple comparisons were controlled at the cluster level using AFNI's 3dClustSim with spatial autocorrelation parameters estimated via 3dLocalACF. This non-stationary cluster-extent correction was based on 10,000 Monte Carlo simulations and yielded a minimum cluster size threshold corresponding to a family-wise error (FWE)–corrected $\alpha = 0.05$, resulting in a cluster extent of ≥40 contiguous voxels.

**Functional connectivity**

ROI-to-ROI connectivity effects were evaluated using parametric second-level generalized linear models, with multiple comparisons across all tested connections controlled using the Benjamini–Hochberg false discovery rate (FDR) procedure. This approach controls the expected proportion of false-positive connections among those identified as significant and is a standard inference option implemented in the CONN toolbox for hypothesis-driven ROI-to-ROI analyses.

**Behavioral**

Group differences in behavioral and questionnaire measures were evaluated using mixed repeated-measures ANOVA with Group (active vs. sham) as a between-subjects factor and Time as a within-subjects factor. When post-hoc pairwise comparisons were performed, multiple comparisons were controlled using false discovery rate (FDR) correction (Benjamini–Hochberg), limiting the expected proportion of false-positive findings across the set of behavioral contrasts.

**Exploratory analyses**

In addition to the primary and secondary hypothesis–driven analyses, exploratory analyses were conducted to examine additional associations among neural, behavioral, and stimulation-related measures. These analyses were not corrected for multiple comparisons, and uncorrected p-values are reported.

**References**


1. Bell, C.C., DSM-IV: diagnostic and statistical manual of mental disorders. Jama, 1994. **272**(10): p. 828-829.
2. Sheehan, D.V., et al., The Mini-International Neuropsychiatric Interview (MINI): the development and validation of a structured diagnostic psychiatric interview for DSM-IV and ICD-10. J clin psychiatry, 1998. **59**(Suppl 20): p. 22-33.
3. Franken, I.H., V.M. Hendriks, and W. van den Brink, Initial validation of two opiate craving questionnaires: the Obsessive Compulsive Drug Use Scale and the Desires for Drug Questionnaire. Addictive behaviors, 2002. **27**(5): p. 675-685.
4. Ekhtiari, H., et al., Methamphetamine and Opioid Cue Database (MOCD): development and validation. Drug and alcohol dependence, 2020. **209**: p. 107941.
